# Signet ring cell histology is an independent predictor of poor prognosis in gastric adenocarcinoma: A population-based analysis

**DOI:** 10.1101/2023.08.06.23293702

**Authors:** Zheyu Huang, Chao Chen, Jianglong Han, Yuxuan Wei, Ruyan Chen, Haiyu Deng, Tingting Jian, Wenmin Liu, Zhenming Fu

## Abstract

**Background:** To test the hypothesis that signet ring cell (SRC) histology is an independent predictor of poor prognosis in gastric adenocarcinoma regardless of tumoral clinical presentation.

**Methods:** We conducted a population-based study to examining the prognostic factors of these two histological subtypes of gastric cancer using data of gastric cancer patients from the Surveillance, Epidemiology, and End Results (SEER) registry between January 2004 and December 2020. Univariate and multivariate Cox regression, and propensity score matching (PSM) models were used to investigate the association between clinical characteristics and prognosis and to calculated hazard ratios (HRs), and corresponding 95% confidence intervals (95% CI).

**Results:** Among a total of 38,336 patients, there were 7,979 SRC and 30,357 non-SRC gastric cancer patients. At presentation, SRC significantly differs from non-SRC patients in the distribution of age, sex, race, primary site and stage. Overall, SRC patients confers worse overall survival (OS: HR = 1.21, 95% CI: 1.17-1.24) and cancer-specific survival (CSS: HR = 1.27, 95% CI: 1.23-1.31) than non-SRC patients. Compared with non-SRC gastric cancer, although stage I SRC has overall better survival (mOS: 90 *vs* 68 months, *P* < 0.001), however, this better survival of SRC was mainly driven by younger age at diagnosis. After adjusted for age at diagnosis as a continuous variable, early stage SRC patients even has a higher risk of mortality (HR = 1.13, 95% CI: 1.03-1.23 and HR = 1.26, 95% CI: 1.13-1.40 for AJCC stage I and II, respectively). While at advanced stages (stage IV), SRC directly confers worse prognosis and has poorer responses to chemotherapy (*P_-heterogeneity_* < 0.001) in either patients with negative (*P _-heterogeneity_* = 0.009) or positive peritoneal cytology (*P_-heterogeneity_* = 0.055).

**Conclusion:** After adjustment of age, SRC confers worse prognosis at all stages. Our study indicates, stage for stage, the SRC histology per se conveys additional risk of mortality. The results support the concept that SRC is a distinct subtype of gastric adenocarcinoma and SRC histology is an independent predictor of poor prognosis for gastric cancer.

## Introduction

Gastric cancer is the fifth most diagnosed cancer and the fourth cause of cancer death worldwide[1]. Most gastric cancer are adenocarcinoma[2], and among them 10-30% are signet ring cell carcinoma (SRC)[3]. Despite worldwide decline in incidence and mortality over the past 5 decades, gastric cancer remains the third leading cause of cancer-related death[4]. However, the incidence of gastric cancer increased in populations younger than 40 years in several countries[5].

SRC gastric cancer is a histologic diagnosis based on microscopic characteristics[6]. It has long been thought to be an aggressive subtype of gastric cancer with a worse prognosis[7, 8]. However, recent results have been controversial. Some reported that SRC histology is an independent predictor of poor prognosis in gastric cancer regardless of tumoral clinical presentation[9]. The other found that when taking extent of disease at presentation into account, SRC histology per se does not portend a worse prognosis[10]. Nevertheless, most studies reported that SRC and non-SRC gastric cancers were not comparable in the distribution of gender[10–17], age[10–17], tumoral stage[10, 15, 16], and primary sites[10, 14, 16–18], and thus leaded to different prognosis. A majority of studies observed that SRC histology is a poor prognostic factor in advanced gastric cancer[9, 11, 14, 16, 19–22]. But the prognostic value for early-stage disease has been highly controversial. Majority reported SRC has a better outcome, particularly when resected endoscopically[23]. Most studies found a better raw survival for SRC in univariable analysis[11, 12, 16, 19, 20, 24] and Kaplan-Meier survival curves[14–17, 19], some observed no difference[22, 25] or worse survival[9] compared with non-SRC patients. Others found the survival advantage despaired after multivariable adjustment[11, 20, 21].

The key issue of the controversy about the prognosis of SRC gastric cancer lines in that whether the prognostic differences between SRC and non-SRC patients was driven by an intrinsically aggressive biology[9] or by different extent of disease at presentation of [10]. Since all SRCs are by definition poorly differentiated, we hypothesize that SRC histology is an independent predictor of poor prognosis in gastric cancer regardless of tumoral clinical presentation. Therefore, to test the hypothesis, we used the large population-based Surveillance, Epidemiology, and End Result (SEER) database and comprehensively investigated the association of clinicopathological features and their interactions in the survival outcomes of gastric cancer.

## Study Population and Methods

### Data Source

The SEER program of the National Cancer Institute (NCI) is an authoritative source of cancer incidence, mortality, and survival in the US, covering approximately 47.9% of the US population[26]. The SEER-17 database recorded cancer survival and therapeutic information from 2000 to 2020. Patients with SRC and non-SRC cancer were selected from the SEER-17 (2004-2020) database using SEER*Stat software 8.4.1.2 (version 8.4.1.2; NCI). Information evaluated from SEER included: patient age, sex, race, primary tumor site, stage at diagnosis, surgery, radiation, and follow-up vital status. Patient consent was not required because the study was a retrospective database research in nature, there was no direct patient contact. Institutional Review Board approval was not required according to our institution policy.

### Study Sample

The SEER-17 records from 2004 to 2020 were correlated with the American Joint Committee on Cancer (AJCC) staging manual (6^th^ edition). SRC is a histologic diagnosis based on microscopic characteristics. The International Classification of Diseases (ICD) code 8490/3 was used to identify patients with SRC carcinoma, whereas code 8140/3, 8144/3, 8145/3, 8211/3, 8260/3, 8480/3 was used for non-SRC. Of 81,282 patients with gastric adenocarcinomas and SRC gastric carcinomas, we excluded the following individuals from the analysis: race unknown (n = 189), without histologically confirmed (n = 425), staging unknown (n = 7,411), surgery status unknown (n = 856), radiation status unknown (n = 593), unknown SEER cause-specific death classification (n = 987), without active follow-up (n = 2,129), with more than two primary tumor (n = 12,666), deaths occurred during the first month of follow up (n = 9,221), and with unknown pathology grade (n = 8,469). This results in a final analytic sample of 38,336 patients (**Figure 1**).

**Figure 1.**
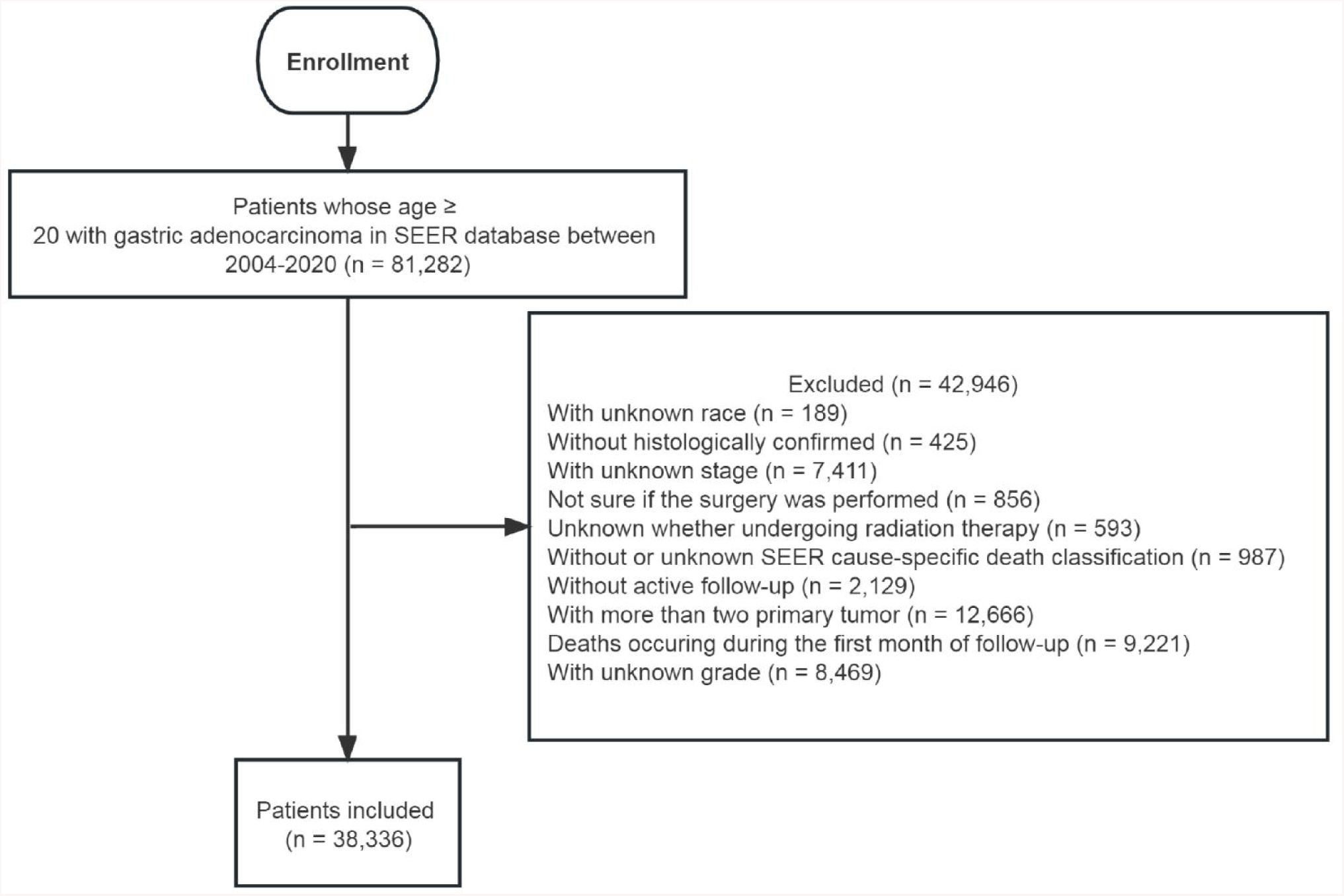
The flowchart of study population selection. Abbreviation: SEER: Surveillance, Epidemiology, End Results

### Statistical Analysis

To compared the baseline and clinic characteristics between SRC and non-SRC, *t* tests and χ2 tests for continuous and categorical variables were used. Propensity score matching (PSM) with nearest neighbor matching was performed in our study to reduce the selection bias and ensure baseline balance between groups, using the calipers equal to 0.05 of the standard deviation from the logit of the propensity score. Variables presented significant in the baseline were selected to generate propensity scores. Surgery was also added to generate propensity scores to make baseline characteristics more balanced between gastric SRC and non-SRC groups. Univariable and multivariable Cox proportional hazard regressions were used to determine the association of mortality with cancer histology type before and after PSM. The results are presented as the hazard ratio (HR) with 95% confidence interval (CI). Likelihood ratio tests for multiplicative interaction were used to compare the models with and without interaction terms to evaluate heterogeneity between SRC and gastric non-SRC cancers. The association and interactions were also examined in various sensitivity analyses to test the robustness. Incidence rates are calculated and age-adjusted to the 2000 US standard population with the age variable recode < 1-year-old. Treatment variables such as surgery, radiotherapy, and chemotherapy were binary. *P-*values for trend tests were derived by entering categorical variables as continuous parameters in the models. *P* values of ≤ 0.05 (2-sided) were considered to be statistically significant. Analyses were conducted using SPSS (version 23, IBM Corp, Armonk, NY, USA) and R version 4.1.2.

## Results

Distributions of characteristics for patients with gastric cancer are presented in **Table 1**. Of the 38,336 patients, 7,979 patients (20.8%) had signet ring cell carcinoma (SRC) and 30,357 patients (79.2%) had non-SRC cancers. Compared with non-SRC patients, SRC patients presented at a younger age (mean age: 59.7 *vs* 66.2 years old; *P* < 0.001), and a smaller proportion of males (50.4% *vs* 67.7%). Non-SRC patients were more frequently white (51.2% *vs* 42.5%) and black (12.1% *vs* 10.7%). SRC patients were more frequently Hispanic ethnicity and Asian/Pacific Islander (API). American Indian/Alaska Native (AIA) patients represented comparable SRC and non-SRC cancers. More SRC located in the overlapping parts of the stomach (26.1% *vs* 15.3%) and less upper stomach (20.5% *vs* 41.9%). In general, SRC cases had significantly higher AJCC stages (*P* < 0.001) and in particular more metastatic disease (stage IV, 47.6% *vs* 39.2%, *P* < 0.001) and more peritoneal metastasis (31.3% *vs* 26.0%, *P* < 0.001). SRC patients received more chemotherapy (69.7% *vs* 61.6%, *P* < 0.001), less radiotherapy (27.3 % *vs* 31.8%, *P* < 0.001), and comparable surgery (57.6% *vs* 57.8%).

**Table 1.**
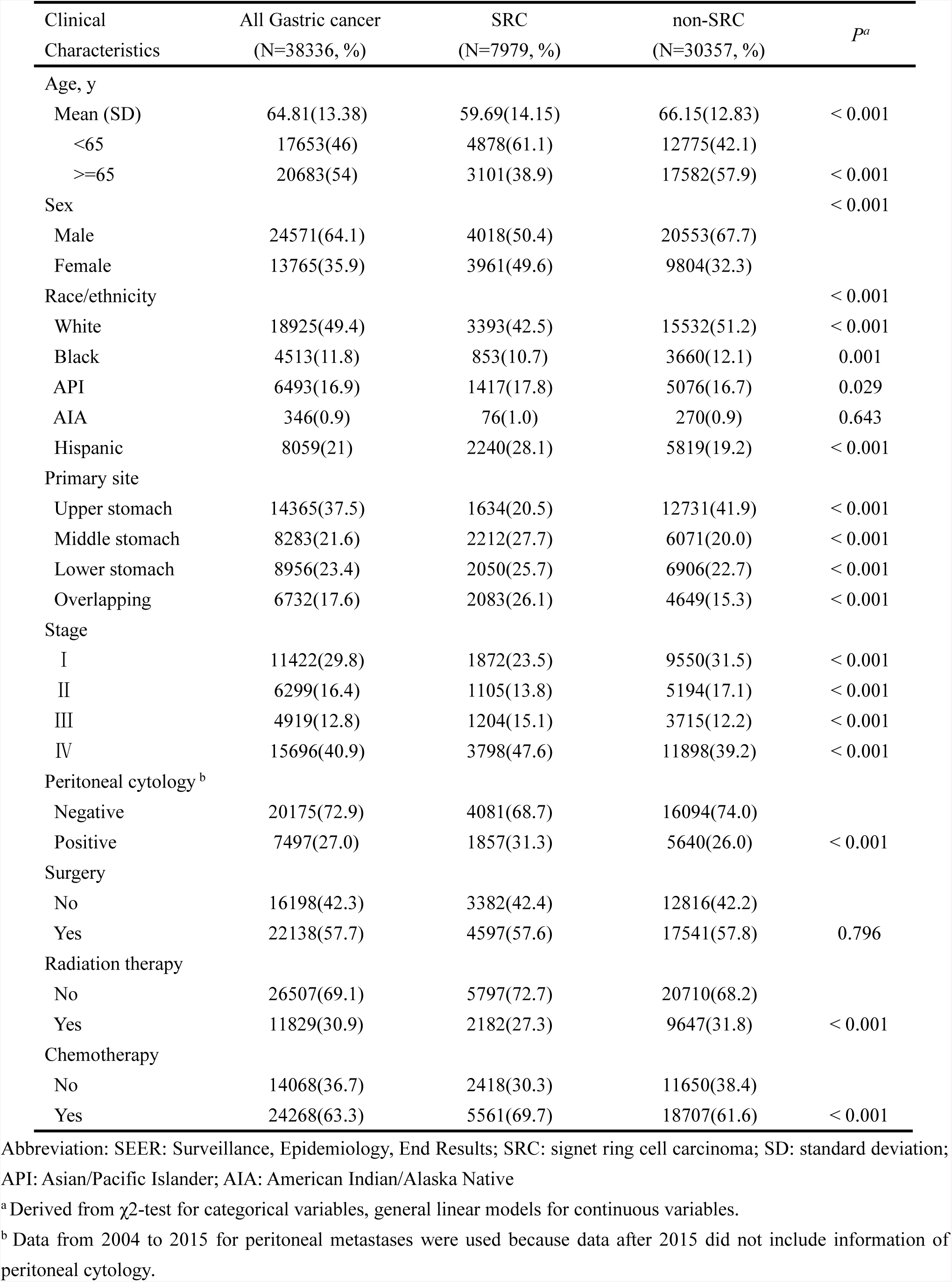
Baseline characteristics of gastric cancer groups, SEER 2004-2020.

| Clinical Characteristics | All Gastric cancer<br>(N=38336, %) | SRC<br>(N=7979, %) | non-SRC<br>(N=30357, %) | *P*<sup>a</sup> |
| --- | --- | --- | --- | --- |
| Age, y |  |  |  |  |
| Mean (SD) | 64.81(13.38) | 59.69(14.15) | 66.15(12.83) | < 0.001 |
| <65 | 17653(46) | 4878(61.1) | 12775(42.1) |  |
| ≥65 | 20683(54) | 3101(38.9) | 17582(57.9) | < 0.001 |
| Sex |  |  |  | < 0.001 |
| Male | 24571(64.1) | 4018(50.4) | 20553(67.7) |  |
| Female | 13765(35.9) | 3961(49.6) | 9804(32.3) |  |
| Race/ethnicity |  |  |  | < 0.001 |
| White | 18925(49.4) | 3393(42.5) | 15532(51.2) | < 0.001 |
| Black | 4513(11.8) | 853(10.7) | 3660(12.1) | 0.001 |
| API | 6493(16.9) | 1417(17.8) | 5076(16.7) | 0.029 |
| AIA | 346(0.9) | 76(1.0) | 270(0.9) | 0.643 |
| Hispanic | 8059(21) | 2240(28.1) | 5819(19.2) | < 0.001 |
| Primary site |  |  |  |  |
| Upper stomach | 14365(37.5) | 1634(20.5) | 12731(41.9) | < 0.001 |
| Middle stomach | 8283(21.6) | 2212(27.7) | 6071(20.0) | < 0.001 |
| Lower stomach | 8956(23.4) | 2050(25.7) | 6906(22.7) | < 0.001 |
| Overlapping | 6732(17.6) | 2083(26.1) | 4649(15.3) | < 0.001 |
| Stage |  |  |  |  |
| I | 11422(29.8) | 1872(23.5) | 9550(31.5) | < 0.001 |
| II | 6299(16.4) | 1105(13.8) | 5194(17.1) | < 0.001 |
| III | 4919(12.8) | 1204(15.1) | 3715(12.2) | < 0.001 |
| IV | 15696(40.9) | 3798(47.6) | 11898(39.2) | < 0.001 |
| Peritoneal cytology<sup>b</sup> |  |  |  |  |
| Negative | 20175(72.9) | 4081(68.7) | 16094(74.0) |  |
| Positive | 7497(27.0) | 1857(31.3) | 5640(26.0) | < 0.001 |
| Surgery |  |  |  |  |
| No | 16198(42.3) | 3382(42.4) | 12816(42.2) |  |
| Yes | 22138(57.7) | 4597(57.6) | 17541(57.8) | 0.796 |
| Radiation therapy |  |  |  |  |
| No | 26507(69.1) | 5797(72.7) | 20710(68.2) |  |
| Yes | 11829(30.9) | 2182(27.3) | 9647(31.8) | < 0.001 |
| Chemotherapy |  |  |  |  |
| No | 14068(36.7) | 2418(30.3) | 11650(38.4) |  |
| Yes | 24268(63.3) | 5561(69.7) | 18707(61.6) | < 0.001 |

Abbreviation: SEER: Surveillance, Epidemiology, End Results; SRC: signet ring cell carcinoma; SD: standard deviation; API: Asian/Pacific Islander; AIA: American Indian/Alaska Native
<sup>a</sup> Derived from $\chi^2$ -test for categorical variables, general linear models for continuous variables.
<sup>b</sup> Data from 2004 to 2015 for peritoneal metastases were used because data after 2015 did not include information of peritoneal cytology.

**Table 2** summarized the predictive factors for mortality of all gastric adenocarcinoma patients. We found older age, black and AIA race, SRC histology, overlapping primary site, higher AJCC stages (**Table 2**) and peritoneal metastasis (**Table S2**) were independent risk factors associated with remarkably worse OS and CSS (all *Ps* < 0.05). In addition, surgical operation, radiation therapy and chemotherapy were found to be strongly associated with better survival (*Ps* < 0.01). As expected, we also found that gastric adenocarcinoma located in the upper stomach had worse survival than those located in the middle or lower stomach (*Ps* < 0.001), while Asian/Pacific Islander (API) patients carried the best survival among all ethnical groups. After PSM, all selected variables were balanced between two groups (**Table 3**). However, there were still significant differences in the OS (mOS: 29.8 *vs* 35.6 months, *P* < 0.01) and CSS (mCSS: 23.2 *vs* 27.0 months, *P* < 0.01) for gastric SRC and non-SRC.

**Table 2.** Association between clinical characteristics with mortality for patients with gastric adenocarcinoma, SEER 2004-2020.

|  |  |  | Overall Survival |  |  |  |  |  | Cancer-specific Survival |  |  |  |  |  |
| --- | --- | --- | --- | --- | --- | --- | --- | --- | --- | --- | --- | --- | --- | --- |
| Clinical Characteristics | N | mOS | Univariable |  |  | Multivariable<sup>a</sup> |  |  | Univariable |  |  | Multivariable<sup>b</sup> |  |  |
|  |  |  | HR | 95% CI | P | HR | 95% CI | P | HR | 95% CI | P | HR | 95% CI | P |
| Age | 38336 | 18 | 1.01 | 1.01-1.01 | < 0.001 | / | / | / | 1.00 | 1.00-1.00 | < 0.001 | / | / | / |
| Age, y |  |  |  |  |  |  |  |  |  |  |  |  |  |  |
| <65 | 17653 | 19 | 1.0 | 1.0 |  | 1.0 | 1.0 |  | 1.0 | 1.0 |  |  | / |  |
| >=65 | 20683 | 18 | 1.15 | 1.12-1.17 | < 0.001 | 1.3 | 1.27-1.34 | < 0.001 | 0.98 | 0.96-1 | 0.115 | / | / | / |
| Histology |  |  |  |  |  |  |  |  |  |  |  |  |  |  |
| Gastric non-SRC (ref.) | 30357 | 19 | 1.0 | 1.0 |  | 1.0 | 1.0 |  | 1.0 | 1.0 |  | 1.0 | 1.0 |  |
| Gastric SRC | 7979 | 15 | 1.16 | 1.13-1.19 | < 0.001 | 1.21 | 1.17-1.24 | < 0.001 | 1.26 | 1.22-1.3 | < 0.001 | 1.27 | 1.23-1.31 | < 0.001 |
| Sex |  |  |  |  |  |  |  |  |  |  |  |  |  |  |
| Male (ref.) | 24571 | 18 | 1.0 | 1.0 |  | / | / |  | 1.0 | 1.0 |  | / | / |  |
| Female | 13765 | 18 | 1 | 0.97-1.02 | 0.792 | / | / | / | 1 | 0.97-1.02 | 0.802 | / | / | / |
| Race/ethnicity |  |  |  |  |  |  |  |  |  |  |  |  |  |  |
| White | 18925 | 17 | 1.0 | 1.0 |  | 1.0 | 1.0 |  | 1.0 | 1.0 |  | 1.0 | 1.0 |  |
| Black | 4513 | 17 | 0.99 | 0.95-1.03 | 0.564 | 1.04 | 1-1.09 | 0.029 | 0.97 | 0.93-1.01 | 0.132 | 1.02 | 0.98-1.06 | 0.511 |
| API | 6493 | 27 | 0.72 | 0.7-0.75 | < 0.001 | 0.79 | 0.76-0.82 | < 0.001 | 0.73 | 0.71-0.76 | < 0.001 | 0.82 | 0.79-0.85 | < 0.001 |
| AIA | 346 | 13 | 1.15 | 1.02-1.3 | 0.021 | 1.23 | 1.09-1.39 | 0.001 | 1.17 | 1.03-1.33 | 0.016 | 1.24 | 1.09-1.41 | 0.025 |
| Hispanic | 8059 | 17 | 0.95 | 0.92-0.98 | 0.001 | 0.92 | 0.89-0.95 | < 0.001 | 1 | 0.96-1.03 | 0.829 | 0.94 | 0.91-0.97 | < 0.001 |
| Primary site |  |  |  |  |  |  |  |  |  |  |  |  |  |  |
| Upper stomach | 14365 | 17 | 1.0 | 1.0 |  | 1.0 | 1.0 |  | 1.0 | 1.0 |  | 1.0 | 1.0 |  |
| Middle stomach | 8283 | 23 | 0.82 | 0.79-0.84 | < 0.001 | 0.96 | 0.93-0.99 | 0.021 | 0.79 | 0.77-0.82 | < 0.001 | 0.94 | 0.91-0.98 | 0.003 |
| Lower stomach | 8956 | 24 | 0.8 | 0.78-0.83 | < 0.001 | 1 | 0.97-1.04 | 0.892 | 0.77 | 0.74-0.79 | < 0.001 | 0.99 | 0.95-1.03 | 0.52 |
| Overlapping | 6732 | 12 | 1.27 | 1.22-1.31 | < 0.001 | 1.15 | 1.11-1.19 | < 0.001 | 1.29 | 1.25-1.34 | < 0.001 | 1.14 | 1.1-1.18 | < 0.001 |
| Stage |  |  |  |  |  |  |  |  |  |  |  |  |  |  |
| I | 11422 | 70 | 1.0 | 1.0 |  | 1.0 | 1.0 |  | 1.0 | 1.0 |  | 1.0 | 1.0 |  |
| II | 6299 | 34 | 1.41 | 1.35-1.46 | < 0.001 | 1.87 | 1.8-1.95 | < 0.001 | 1.76 | 1.68-1.85 | < 0.001 | 2.31 | 2.2-2.42 | < 0.001 |
| III | 4919 | 20 | 2.04 | 1.96-2.13 | < 0.001 | 2.81 | 2.7-2.94 | < 0.001 | 2.73 | 2.61-2.86 | < 0.001 | 3.67 | 3.49-3.85 | < 0.001 |
| IV | 15696 | 9 | 4.58 | 4.43-4.72 | < 0.001 | 3.83 | 3.69-3.98 | < 0.001 | 6.33 | 6.1-6.57 | < 0.001 | 4.99 | 4.78-5.21 |  |
| *P<sub>trend</sub>* |  |  | 1.66 | 1.64-1.68 | < 0.001 | 1.54 | 1.52-1.56 | < 0.001 | 1.84 | 1.82-1.86 | < 0.001 | 1.64 | 1.62-1.67 | < 0.001 |
| Peritoneal cytology<sup>c</sup> |  |  |  |  |  |  |  |  |  |  |  |  |  |  |
| Negative | 20175 | 28 | 1.0 | 1.0 |  |  |  |  | 1.0 | 1.0 |  |  |  |  |
| Positive | 7497 | 7 | 3.4 | 3.3-3.5 | < 0.001 |  |  |  | 3.82 | 3.71-3.94 | < 0.001 |  |  |  |
| Surgery |  |  |  |  |  |  |  |  |  |  |  |  |  |  |
| No (ref.) | 16198 | 9 | 1.0 | 1.0 |  | 1.0 | 1.0 |  | 1.0 | 1.0 |  | 1.0 | 1.0 |  |
| Yes | 22138 | 42 | 0.26 | 0.25-0.27 | < 0.001 | 0.35 | 0.34-0.36 | < 0.001 | 0.23 | 0.23-0.24 | < 0.001 | 0.34 | 0.33-0.35 | < 0.001 |
| Radiation therapy |  |  |  |  |  |  |  |  |  |  |  |  |  |  |
| No (ref.) | 26507 | 17 | 1.0 | 1.0 |  | 1.0 | 1.0 |  | 1.0 | 1.0 |  | 1.0 | 1.0 |  |
| Yes | 11829 | 22 | 0.87 | 0.85-0.89 | < 0.001 | 1.09 | 1.06-1.12 | < 0.001 | 0.89 | 0.86-0.91 | < 0.001 | 1.1 | 1.07-1.14 | 0.061 |
| Chemotherapy |  |  |  |  |  |  |  |  |  |  |  |  |  |  |
| No (ref.) | 14068 | 21 | 1.0 | 1.0 |  | 1.0 | 1.0 |  | 1.0 | 1.0 |  | 1.0 | 1.0 |  |
| Yes | 24268 | 18 | 1.13 | 1.11-1.16 | < 0.001 | 0.58 | 0.56-0.59 | < 0.001 | 1.3 | 1.26-1.34 | < 0.001 | 0.58 | 0.56-0.6 | < 0.001 |

Abbreviation: SEER: Surveillance, Epidemiology, End Results; ref.: referent; HR: hazards ratio; CI: Confidence interval; mOS: median overall survival; SRC: signet ring cell carcinoma; API: Asian/Pacific Islander; AIA: American Indian/Alaska Native
<sup>a</sup> Variables included in multivariate analysis for overall survival were age ( $\geq 65$ , $< 65$ years old), primary site, histology, race, stage, surgery, radiation therapy and chemotherapy.
<sup>b</sup> Variables included in multivariate analysis for cause-specific survival were primary site, histology, race, stage, surgery, radiation therapy and chemotherapy.
<sup>c</sup> Data from 2004 to 2015 for peritoneal metastases were used because data after 2015 did not include information of peritoneal cytology.

**Table 3.**
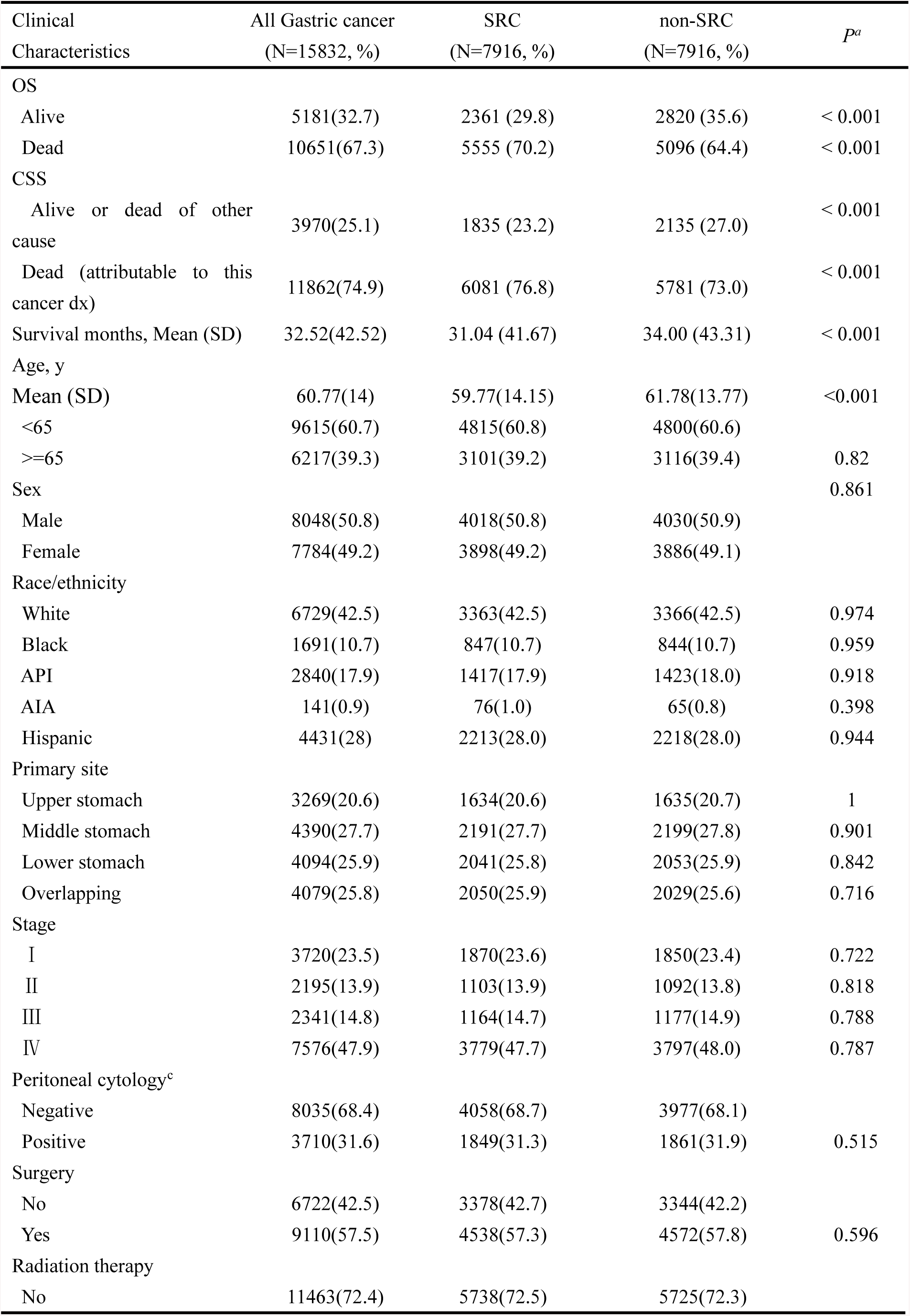
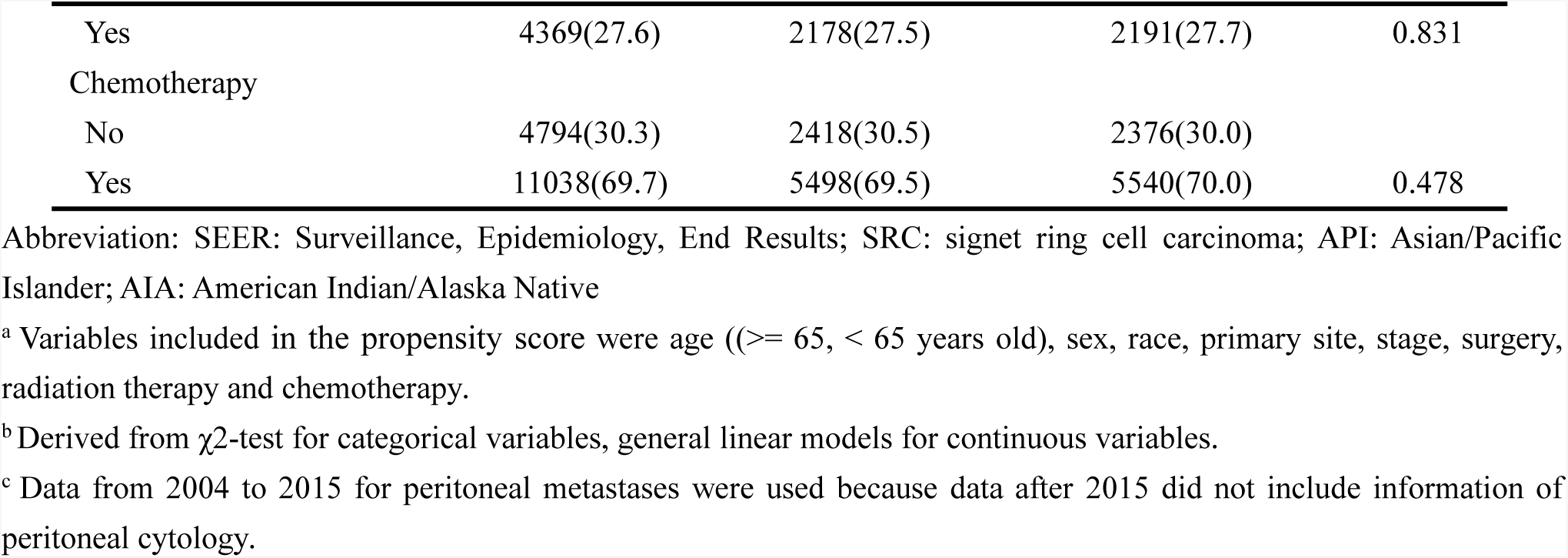
Baseline characteristics of gastric cancer (signet ring cell carcinoma and non-signet ring cell carcinoma) groups after propensity score matching^a^, SEER 2004-2020.

Interaction tests were conducted to assess the heterogeneity in the effects of clinical factors on the survival between SRC and non-SRC patients (**Table 4**). There was no effect modification on the association between OS of SRC and that of non-SRC patients by age, sex, race, peritoneal metastasis and whether received surgery with all interaction coefficients (**Table 4, S4.1-4.4**). However, there were significant heterogeneity in the association of OS with AJCC stage (*P_-interaction_* < 0.001) and chemotherapy (*P_-interaction_* < 0.001) between SRC and non-SRC patients (**Table S4.4**). After PSM, similar heterogenous effects were found on the cancer specific survivals for AJCC stage (*P_-interaction_* = 0.007) and chemotherapy (*P_-interaction_* < 0.001) between SRC and non-SRC patients **(Table 4)**. In some different ways, we find there were also significant heterogeneity in the association of OS with primary site and radiation therapy (**Table S4.1-S4.4**).

**Table 4.** Heterogeneity in the association of survival with clinical characteristics between patients with gastric signet ring cell carcinoma and non-signet ring cell carcinoma after propensity score matching^a^, SEER 2004-2020.

| Clinical characteristics | N, non-SRC (ref.) | N, SRC | OS |  |  |  |  | *P*<sub>heterogeneity</sub> | CSS |  |  |  |  | *P*<sub>heterogeneity</sub> |
| --- | --- | --- | --- | --- | --- | --- | --- | --- | --- | --- | --- | --- | --- | --- |
|  |  |  | mOS (non-SRC) | mOS (SRC) | HR | 95% CI | *P* |  | mCSS (non-SRC) | mCSS (SRC) | HR | 95% CI | *P* |  |
| Age (continuous variable, Mean (SD)) | 61.8 (13.8) | 59.8 (14.2) | 18 | 15 |  |  |  | 0.762 | 20 | 16 |  |  |  | 0.16 |
| Sex |  |  |  |  |  |  |  | 0.275 |  |  |  |  |  | 0.205 |
| Male | 4030 | 4018 | 19 | 15 | 1.18 | 1.12-1.24 | < 0.001 |  | 20 | 15 | 1.21 | 1.14-1.27 | < 0.001 |  |
| Female | 3886 | 3898 | 18 | 15 | 1.14 | 1.08-1.2 | < 0.001 |  | 19 | 16 | 1.16 | 1.1-1.23 | < 0.001 |  |
| Race/ethnicity |  |  |  |  |  |  |  | 0.826 |  |  |  |  |  | 0.765 |
| White | 3366 | 3363 | 17 | 14 | 1.16 | 1.1-1.23 | < 0.001 |  | 19 | 15 | 1.18 | 1.11-1.25 | < 0.001 |  |
| Black | 844 | 847 | 19 | 15 | 1.16 | 1.04-1.29 | 0.01 |  | 20 | 17 | 1.18 | 1.05-1.33 | 0.007 |  |
| API | 1423 | 1417 | 24 | 20 | 1.11 | 1.02-1.22 | 0.022 |  | 27 | 21 | 1.13 | 1.02-1.24 | 0.014 |  |
| AIA | 65 | 76 | 12 | 15 | 1.11 | 0.75-1.64 | 0.59 |  | 12 | 15 | 1.14 | 0.75-1.73 | 0.532 |  |
| Hispanic | 2218 | 2213 | 16 | 13 | 1.16 | 1.08-1.24 | < 0.001 |  | 17 | 14 | 1.2 | 1.11-1.29 | < 0.001 |  |
| Primary site |  |  |  |  |  |  |  | 0.709 |  |  |  |  |  | 0.901 |
| Upper stomach | 1635 | 1634 | 17 | 13 | 1.19 | 1.1-1.28 | < 0.001 |  | 18 | 14 | 1.21 | 1.12-1.31 | < 0.001 |  |
| Middle stomach | 2199 | 2191 | 22 | 18 | 1.14 | 1.06-1.23 | < 0.001 |  | 25 | 19 | 1.18 | 1.09-1.27 | < 0.001 |  |
| Lower stomach | 2053 | 2041 | 24 | 20 | 1.17 | 1.09-1.26 | < 0.001 |  | 28 | 22 | 1.21 | 1.11-1.31 | < 0.001 |  |
| Overlapping | 2029 | 2050 | 13 | 11 | 1.16 | 1.08-1.24 | < 0.001 |  | 13 | 12 | 1.18 | 1.1-1.26 | < 0.001 |  |
| Stage |  |  |  |  |  |  |  | 0.007 |  |  |  |  |  | 0.791 |
| I | 1850 | 1870 | 83 | 90 | 1.13 | 1.03-1.23 | 0.012 |  |  |  | 1.27 | 1.14-1.42 | < 0.001 |  |
| II | 1092 | 1103 | 41 | 33 | 1.26 | 1.13-1.4 | < 0.001 |  | 51 | 36 | 1.29 | 1.15-1.45 | < 0.001 |  |
| III | 1177 | 1164 | 23 | 18 | 1.3 | 1.18-1.42 | < 0.001 |  | 26 | 20 | 1.32 | 1.19-1.45 | < 0.001 |  |
| IV | 3797 | 3779 | 10 | 8 | 1.19 | 1.13-1.25 | < 0.001 |  | 10 | 8 | 1.2 | 1.14-1.26 | < 0.001 |  |
| Peritoneal cytology<sup>b</sup> |  |  |  |  |  |  |  | 0.107 |  |  |  |  |  | 0.577 |
| Negative | 3977 | 4058 | 29 | 23 | 1.17 | 1.11-1.23 | < 0.001 |  | 35 | 26 | 1.22 | 1.15-1.29 | < 0.001 |  |
| Positive | 1861 | 1849 | 8 | 7 | 1.19 | 1.11-1.27 | < 0.001 |  | 8 | 7 | 1.19 | 1.11-1.27 | < 0.001 |  |
| Surgery |  |  |  |  |  |  |  | 0.122 |  |  |  |  |  | 0.712 |
| No | 3344 | 3378 | 9 | 8 | 1.18 | 1.12-1.24 | < 0.001 |  | 9 | 8 | 1.19 | 1.13-1.25 | < 0.001 |  |
| Yes | 4572 | 4538 | 38 | 31 | 1.18 | 1.12-1.24 | < 0.001 |  | 49 | 35 | 1.23 | 1.16-1.3 | < 0.001 |  |
| Radiation therapy |  |  |  |  |  |  |  | 0.164 |  |  |  |  |  | 0.258 |
| No | 5725 | 5738 | 16 | 13 | 1.13 | 1.08-1.17 | < 0.001 |  | 17 | 14 | 1.15 | 1.1-1.21 | < 0.001 |  |
| Yes | 2191 | 2178 | 24 | 20 | 1.21 | 1.13-1.29 | < 0.001 |  | 28 | 21 | 1.23 | 1.14-1.32 | < 0.001 |  |
| Chemotherapy |  |  |  |  |  |  |  | < 0.001 |  |  |  |  |  | 0.028 |
| No | 2376 | 2418 | 18 | 15 | 1.08 | 1.01-1.16 | 0.017 |  | 23 | 18 | 1.14 | 1.06-1.23 | < 0.001 |  |
| Yes | 5540 | 5498 | 18 | 15 | 1.19 | 1.14-1.24 | < 0.001 |  | 19 | 15 | 1.21 | 1.15-1.26 | < 0.001 |  |

Abbreviation: SEER: Surveillance, Epidemiology, End Results; ref.: referent; HR: hazards ratio; CI: Confidence interval; mOS: median overall survival; mCSS: median cause-specific survival; SRC: signet ring cell carcinoma;
API: Asian/Pacific Islander; AIA: American Indian/Alaska Native
<sup>a</sup> Adjusted for propensity score which included age ( $\geq 65$ , $< 65$ years old), sex, race, primary site, stage, surgery, radiation therapy and chemotherapy) and age (as continuous variable).<sup>b</sup> Data from 2004 to 2015 for peritoneal metastases were used because data after 2015 did not include information of peritoneal cytology.

**Table 5** shows the heterogeneities between SRC and non-SRC patients regarding mortality risks modified by stage and treatment. Compared with non-SRC gastric adenocarcinoma patients, at early stages (stage I+II), surgery significantly more effective for SRC patients with better reduced mortality risk (HR = 1.14 *vs* 1.27) with interaction coefficients (*P_-heterogeneity_* < 0.001). The same heterogenous effect of surgery were found in both stage I disease (*P_-heterogeneity_ <* 0.001) and stage II disease (*P_-heterogeneity_* = 0.002), although effect appeared more prominent in stage I disease. Without surgery, SRC patients had 25% higher mortality than non-SRC patients (HR = 1.25, 95% CI: 1.12-1.39), but when received surgery, the mortality for SRC patients became only 9% higher (HR = 1.07, 95% CI: 0.97-1.18). On the other hand, higher mortality was observed for late stage (stage IV) SRC diseases (HR = 1.2 *vs* 1.03, *P_-heterogeneity_* = 0.012) regardless of peritoneal metastasis (*P_-heterogeneity_* = 0.055) or not (*P_-heterogeneity_* = 0.009) in terms of chemotherapy. In general, poorer chemotherapy responses were observed for SRC than non-SRC cancers regardless of stage. When received chemotherapy, SRC patients had a 17% higher mortality than non-SRC patients in stage I and stage II patients (*P_-heterogeneity_* < 0.001 and *P_-heterogeneity_* = 0.431, respectively). For stage III disease, although the heterogeneity was not significant, numerically, SRC patients derived less benefit from chemotherapy (HR = 1.31 *vs* 1.19) than non-SRC patients with gastric cancer.

**Table 5.** Heterogeneity in the association of treatment modality with overall survival by stages between SRC and non-SRC gastric cancers, SEER 2004-2020.

| Stage | non-SRC | SRC | HR (95% CI) | non-SRC | SRC | HR (95% CI) | *P*<sub>heterogeneity</sub> |
| --- | --- | --- | --- | --- | --- | --- | --- |
|  | n | n |  | n | n |  |  |
|  | Surgery (no) |  |  | Surgery (yes) |  |  |  |
| I | 2335 | 476 | 1.25(1.12-1.39) | 7215 | 1396 | 1.07(0.97-1.18) | < 0.001 |
| II | 855 | 189 | 1.37(1.15-1.62) | 4339 | 916 | 1.19(1.09-1.31) | 0.002 |
| III | 584 | 160 | 1.12(0.92-1.36) | 3131 | 1044 | 1.26(1.16-1.37) | 0.734 |
| IV | 9042 | 2557 | 1.17(1.12-1.23) | 2856 | 1241 | 1.22(1.14-1.32) | 0.642 |
| -with peritoneal cytology negative | 1316 | 327 | 1.11(0.98-1.27) | 1277 | 554 | 1.28(1.15-1.42) | 0.248 |
| -with peritoneal cytology positive | 4640 | 1434 | 1.13(1.06-1.2) | 1000 | 422 | 1.22(1.07-1.38) | 0.29 |
|  | Chemotherapy (no) |  |  | Chemotherapy (yes) |  |  |  |
| I | 6139 | 1065 | 1.14(1.03-1.26) | 3411 | 807 | 1.40(1.26-1.56) | < 0.001 |
| II | 1311 | 217 | 1.37(1.16-1.6) | 3883 | 888 | 1.25(1.13-1.37) | 0.431 |
| III | 1086 | 254 | 1.19(1.03-1.39) | 2629 | 950 | 1.31(1.2-1.43) | 0.179 |
| IV | 3114 | 882 | 1.03(0.95-1.11) | 8784 | 2916 | 1.20(1.14-1.26) | < 0.001 |
| -with peritoneal cytology negative | 754 | 226 | 0.98(0.84-1.15) | 1839 | 657 | 1.23(1.12-1.36) | 0.009 |
| -with peritoneal cytology positive | 1659 | 479 | 1.05(0.94-1.17) | 3981 | 1378 | 1.18(1.11-1.27) | 0.055 |

Abbreviations: SEER: Surveillance, Epidemiology; SRC: Signet ring cell carcinoma; HR: hazards ratio; CI: Confidence interval
Adjusted for age (as continuous variable), sex, race and primary site.

In the univariable analyses, we found that, overall compared with non SRC patients, stage I SRC patients had better survival (90 *vs* 68 months, *P* < 0.001), stage II SRC had similar (33 *vs* 34 months, *P* = 0.245), but stage III (19 *vs* 24 months, *P* = 0.001) and stage IV (8 *vs* 9 months, *P* < 0.001) both had worse survival (**Figure 2**). However, in any multivariable models with adjustment with age as a continuous variable, we found SRC had a worse prognosis than non-SRC regardless of stage (**Table 4, S4.1-S4.4)**. But once we did not adjust age, we found SRC had a better prognosis than non-SRC in stage I (**Table S4.5-S4.9**). It may suggest that SRC may have a better prognosis before PSM in stage I because SRC patients were much younger (62.6 *vs* 69.8 years old, *P* < 0.001).

**Figure 2.**
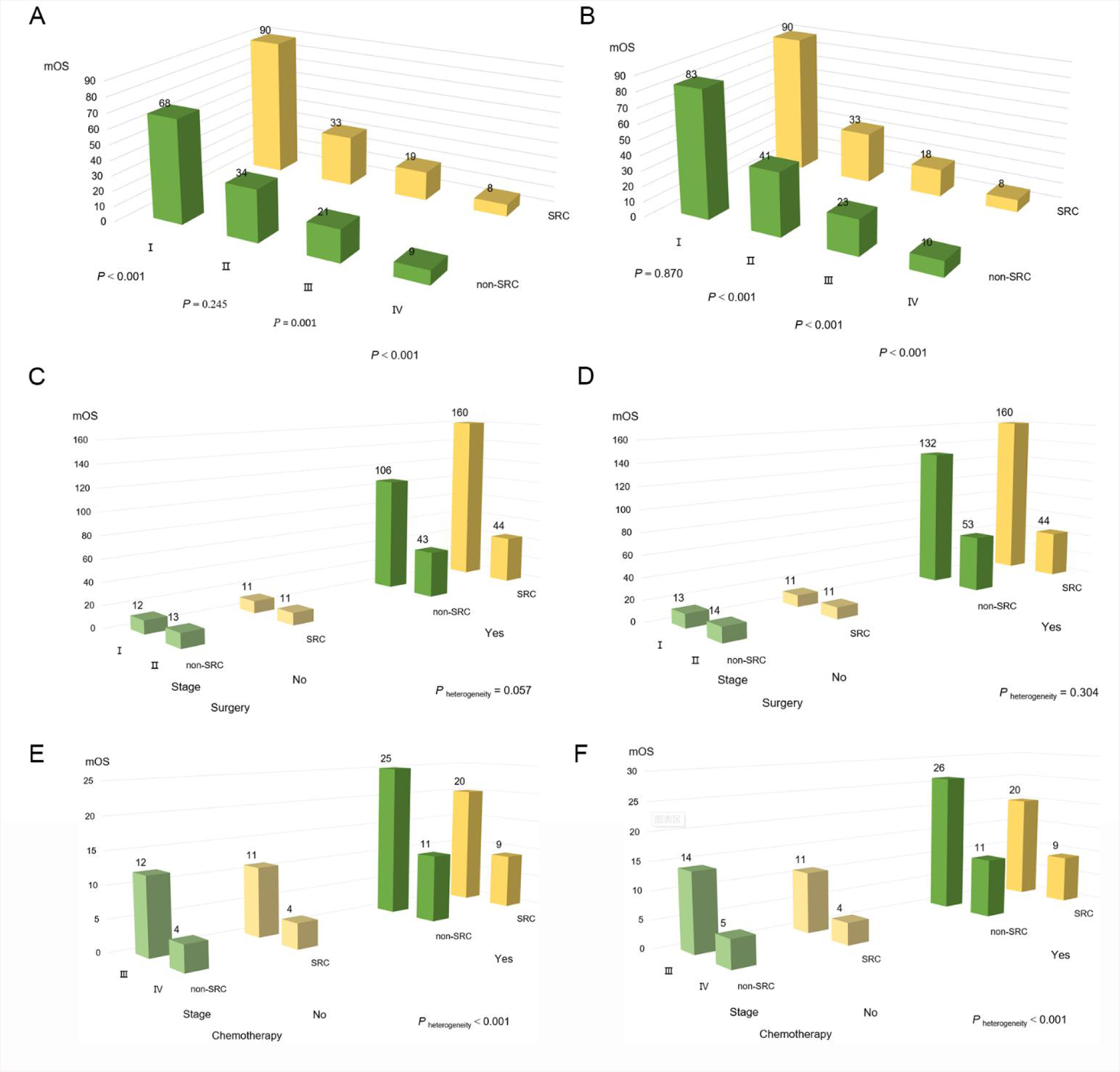
The months of survival between SRC and non-SRC gastric cancers。 (A) all American Joint Committee on Cancer, 6th edition (AJCC) stages before PSM, (B) all stages after PSM, (C)a AJCC stage I and 2 tumors by surgery before PSM, (D)aAJCC stage I and 2 tumors by surgery after PSM; (E)a AJCC stage 3 and 4 tumors by chemotherapy before PSM, and (F)a AJCC stage 3 and 4 tumors by chemotherapy after PSM. a Adjusted for age (as continuous variable), sex, race, primary site, stage, surgery, radiation therapy and chemotherapy.

**Figure 3.**
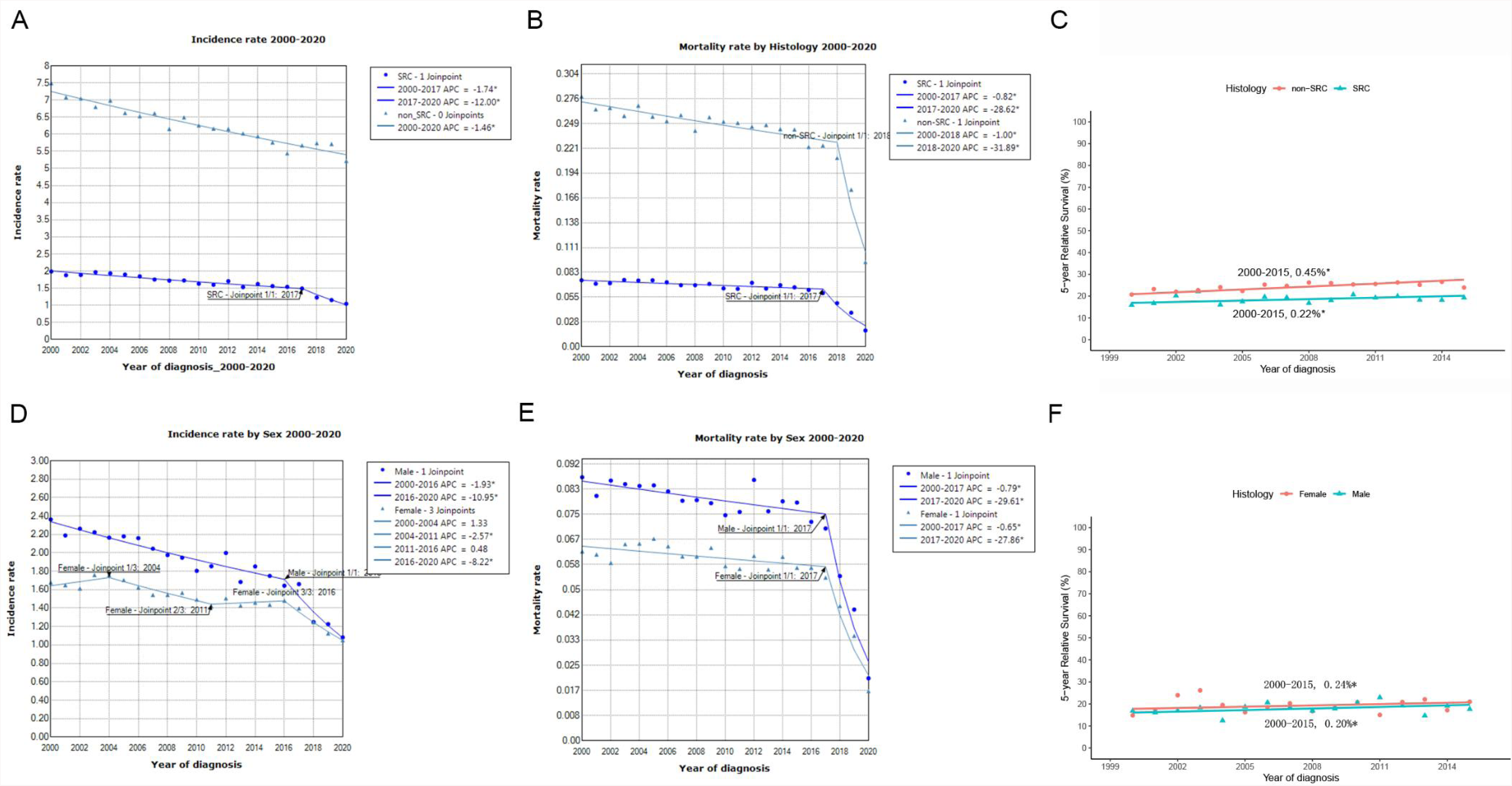
Trends of gastric signet ring cell carcinoma and non-signet ring cell carcinoma, SEER 2000-2020. (A). Incidence rate of 2000-2020. (B). Mortality rate of 2000-2020. (C).5-years survival rate of 2000-2015. (D). Incidence rate by sex of gastric signet ring cell carcinoma, 2000-2020. (B). Mortality rate by sex of gastric signet ring cell carcinoma, 2000-2020. (C).5-years survival rate by sex of gastric signet ring cell carcinoma, 2000-2015.

**Table 6** further demonstrated the overwhelming effects of age at diagnosis on the survival of early stage SRC patients. In the univariate analysis without adjustment of any confounders, the survival time (**Figure 2**) of stage I SRC gastric cancer patients seemed significantly better than non-SRC patients (90 *vs* 68 months, HR = 0.87, 95% CI: 0.81-0.93), especially for those patients received surgery (159 *vs* 120 months, HR = 1.15, 95% CI: 1.07-1.23). However, after adjusting only one variable: age at diagnosis, the survival advantage reversed or disappeared for SRC patients compared with non-SRC patients (HR = 0.74, 95% CI: 0.67-0.82), even for those received surgery (HR = 0.98, 95% CI: 0.89-1.08, *P* = 0.717). In addition, in any multivariable models adjusted for age or in PSM models (age included), the survival advantage of stage I SRC over non-SRC gastric cancer patients reversed. For example, in the PSM model, the survival benefit for stage I SRC gastric cancer patients disappeared (HR = 1.01, 95% CI: 0.92-1.10, *P* = 0.870), including those patients received surgery (160 *vs* 132 months HR = 0.91, 95% CI: 0.81-1.03, *P* = 0.132). On the other hand, in models adjusted for all other confounders but not age, the survival of stage I SRC gastric cancer patients still seemed better than non-SRC patients. For example, in the model adjusted for sex, race, primary site, stage, surgery, radiation therapy, and chemotherapy, the survival time of stage I SRC gastric cancer patients seemed significantly better than non-SRC patients (HR = 0.92, 95% CI: 0.85-0.98), in particular for those patients received surgery (HR = 0.76, 95% CI: 0.69-0.84). Thus, younger age at diagnosis played an overwhelming effect of on the better survival of early stage SRC patients.

**Table 6.** The effects of age on overall survival in early stage SRC and non-SRC gastric cancers, SEER 2004-2020.

|  |  | Surgery<br>or not | Mean of Age (SD) |  | mOS |  | Model1<br>(Univariable) |  | Model2<sup>a</sup><br>(Multivariable with age) |  | Model3<sup>b</sup><br>(Multivariable without age) |  | Model4<sup>c</sup><br>(Multivariable with all variables) |  |
| --- | --- | --- | --- | --- | --- | --- | --- | --- | --- | --- | --- | --- | --- | --- |
|  |  |  | Non-SRC | SRC | Non-SRC | SRC | HR (95% CI) | P | HR (95% CI) | P | HR (95% CI) | P | HR (95% CI) | P |
| Before<br>PSM | I | All | 69.8 (11.7) | 62.6 (14.0) | 68 | 90 | 0.87 (0.81-0.93) | < **0.001** | 1.15 (1.07-1.23) | < 0.001 | 0.92 (0.85-0.98) | **0.015** | 1.14 (1.06-1.23) | < 0.001 |
|  |  | No | 74.0 (11.3) | 67.8 (14.0) | 12 | 11 | 1.13 (1.02-1.26) | 0.021 | 1.27 (1.14-1.42) | < 0.001 | 1.20 (1.04-1.29) | 0.009 | 1.28 (1.14-1.43) | < 0.001 |
|  |  | Yes | 68.4 (11.5) | 60.8 (13.6) | 106 | 160 | 0.74 (0.67-0.82) | < **0.001** | 0.98 (0.89-1.08) | 0.717 | 0.76 (0.69-0.84) | < **0.001** | 1.07 (0.97-1.18) | 0.184 |
|  | II | All | 66.2 (12.2) | 61.4 (13.3) | 34 | 32 | 1.05 (0.97-1.13) | 0.245 | 1.18 (1.09-1.27) | < 0.001 | 1.17 (1.08-1.27) | < 0.001 | 1.26 (1.16-1.36) | < 0.001 |
|  |  | No | 69.1 (12.1) | 65.7 (13.8) | 13 | 11 | 1.34 (1.14-1.58) | 0.001 | 1.38 (1.17-1.63) | < 0.001 | 1.32 (1.11-1.57) | 0.002 | 1.34 (1.13-1.60) | 0.001 |
|  |  | Yes | 65.6 (12.2) | 60.5 (13.0) | 43 | 44 | 1.01 (0.92-1.10) | 0.888 | 1.13 (1.03-1.24) | 0.009 | 1.12 (1.02-1.23) | 0.017 | 1.23 (1.12-1.35) | < 0.001 |
| After<br>PSM<sup>d</sup> | I | All | 65.5 (13.4) | 62.6 (14.0) | 83 | 90 | 1.01 (0.92-1.10) | 0.870 | 1.14 (1.04-1.25) | 0.006 | 1.07 (0.97-1.17) | 0.171 | 1.16 (1.06-1.27) | 0.002 |
|  |  | No | 70.3 (13.4) | 67.8 (14.0) | 13 | 11 | 1.22 (1.06-1.40) | 0.006 | 1.26 (1.10-1.46) | 0.001 | 1.23 (1.06-1.41) | 0.005 | 1.28 (1.11-1.47) | 0.001 |
|  |  | Yes | 64.0 (13.1) | 60.8 (13.6) | 132 | 160 | 0.91 (0.81-1.03) | 0.135 | 1.03 (0.92-1.17) | 0.585 | 0.92 (0.82-1.04) | 0.175 | 1.06 (0.93-1.19) | 0.388 |
|  | II | All | 63.0 (13.0) | 61.4 (13.3) | 41 | 32 | 1.22 (1.10-1.35) | < 0.001 | 1.27 (1.14-1.41) | < 0.001 | 1.25 (1.13-1.39) | < 0.001 | 1.29 (1.16-1.43) | < 0.001 |
|  |  | No | 67.1 (12.8) | 65.7 (13.8) | 14 | 11 | 1.47 (1.17-1.83) | 0.001 | 1.49 (1.19-1.86) | 0.001 | 1.45 (1.15-1.82) | 0.002 | 1.46 (1.16-1.84) | 0.001 |
|  |  | Yes | 62.3 (12.9) | 60.5 (13.0) | 53 | 44 | 1.18 (1.05-1.33) | 0.007 | 1.23 (1.09-1.38) | 0.001 | 1.20 (1.06-1.35) | 0.003 | 1.23 (1.09-1.39) | 0.001 |

Abbreviation: SEER: Surveillance, Epidemiology; SRC: Signet ring cell carcinoma; HR: hazards ratio; CI: Confidence interval
<sup>a</sup>Adjusted for age (continuous variable).
<sup>b</sup>Adjusted for sex, race, primary site, stage, surgery, radiation therapy, chemotherapy.
<sup>c</sup>Adjusted for age (continuous variable), sex, race, primary site, stage, surgery, radiation therapy, chemotherapy.
<sup>d</sup>Adjusted for age ( $\geq 65$ , $< 65$ years old), sex, race, primary site, stage, surgery, radiation therapy, chemotherapy.

## Discussion

The present study is a comprehensive SEER analysis to use PSM and interaction analysis to examine the relative contribution and heterogeneity of possible risk factors on prognosis of SRC and non-SRC patients. To the best of our knowledge, for the first time, our analyses clearly indicated that SRC histology is an independent predictor of poor prognosis in gastric cancer after taking clinical presentations into consideration.

Different from an earlier SEER analysis[10], our analyses demonstrated SRC was independently associated with worse prognosis both for the early stages and for the advanced stages of the disease whenever adjusted for age (as a continuous variable). Although stage for stage, Taghavi *et al* did not find SRC histology bestow additional risk of mortality[10]. Both analyses share much more commonality than differences. Both studies consistently found SRC significantly differs from non-SRC in extent of disease at presentation in the US. For example: SRC are more often found in the distal part of the stomach, in young and female patients, and present at more advanced TNM stage, with a metastatic stage upon diagnosis in about 50% of patients. And mortality was associated with age, black race, and also tumor grade[10]. Also, both studies found significant heterogenous treatment effects between SRC and non-SRC patients: surgery worked better in early-stage (AJCC stage I or II) gastric SRC than non-SRC patients; And SRC seemed resistant to and chemotherapy at all stages, especially in stage IV disease.

Consistent with most other studies[9, 11, 14, 16, 19–22], we found that advanced SRC had worse survival than non SRC patients. Advanced SRC patients tended to have more peritoneal metastasis and did not response well to chemotherapy[27]. Previous studies account these for its worse survival[24]. However, in our sensitivity analyses including models adjusted for peritoneal metastasis and chemotherapy, the worse survival remained (**Table S2-3**). Thus, our finding pointed to worse biology in addition to more extensive of disease for advanced SRC. For early stage SRC, like most previous findings[11, 12, 14–16, 18–20, 24], we found a better raw survival than non-SRC patients. Also consistent with some previous studies, we found the survival advantage despaired after multivariable adjustment[20, 21]. In the current study, whenever age was adjusted as a continuous variable, the better prognosis of early SRC reversed. Previous studies reported early-stage SRC a better survival than non-SRC partially because they had better prognosis when resected endoscopic[23, 24]. However, in our sensitivity analyses which including only patients received surgery (these patients had an even better survival), once age at diagnosis was adjusted as a continuous variable, the better prognosis of SRC again disappeared. Our results may indicate younger age at diagnosis played a critical role in the better survival of early stage SRC patients.

Previously, three early studies[11, 20, 21] included age as a continuous variable in multivariate analyses. They reported that SRC had a better prognosis than either non-SRC[20, 21] or well/median differentiated gastric cancer[11]. However, two studies[20, 21] included patients before 2004, when the diagnostic criteria for SRC and treatment were still in development. While the other one present data from a large Asian population, but only present survival in Kaplan-Meier survival curves which is expected to be univariable data[11]. Many studies did point out that the observed better survival might be related to the younger age at presentation for SRC patients[12, 16]. One study also found that SRC was independently associated with a dismal prognosis using machine matching (which included age as a categorical variable)[9], although the sample size is small. Another study also found that the survival benefit in early SRC patients was not evident when considering exclusively disease-specific survival or in multivariable analysis[12]. Likewise, we have done many kinds of sensitivity analyses, only after adjusting age of diagnosis as a continuous variable, the survival advantage reversed. In our PSM model (which including age as a categorical variable), the survival benefit only disappeared for stage I SRC gastric cancer patients (HR = 1.01, *P* = 0.870), including those patients received surgery (HR = 0.91, *P* = 0.132). On the other hand, in any multivariable models without adjustment of age, stage I SRC still had a relatively better survival than non-SRC disease. Thus, only adjusting age as a continuous variable can effectively rule out the confounding effect of age on the survival of stage I SRC patients. Using age as a categorial variable apparently, will lose a lot of information in the multivariable adjustments. Apparently, in the current study, younger age at diagnosis (62.6 *vs* 69.8, *P* < 0.001) played an overwhelming role in the better survival of early stage SRC patients.

Therefore, our analyses demonstrated that SRC histology independently conferred worse prognosis. In other words, our results support the concept that SRC is a distinct histology type of gastric adenocarcinoma which *per se* bestow worse prognosis. SRC histology has traditionally been regard as with poor prognosis. Over the years, although the classification of SRC gastric cancer has evolved 5 times, World Health Organization (WHO) pathological definitions described more aggressive features than other gastric adenocarcinomas[28–30]. Before 2010, SRC-GC was classified as a separate specific subtype of GC. In 2010, the SRC-GC was redefined entirely as a subtype of poorly cohesive cells gastric cancer[29]. Alternative classification other than WHO systems such as the Lauren and the Ming classification, also categorized SRC-GC as ‘diffuse/mixed’ and ‘infiltrative’ type carcinoma[7, 31]. As such, all definitions and classifications identify SRC histology as unfavorable subtype. It is thus reasonable to expect SRC is driven by an intrinsically aggressive biology with poor prognosis compared with other gastric adenocarcinomas[9]. Recent laboratory studies also reveal that SRC demonstrates distinct cytological and microenvironment features[32–34]. SRC enriched in abnormally activated cancer-related signaling pathways and an immunosuppressive microenvironment[34]. Another single-cell study also found the immune irresponsiveness of SRC[32]. These studies support that SRC is a distinct entity. The molecular characterization of SRC and gastric cancer will certainly improve our understanding of the disease and improves our ability to predict treatment response to and to develop targeted therapeutics[33].

There are similar SEER studies[10, 19, 35], none of them performed PSM. One study investigated gastric cancer as a whole without histology subgroup analysis[35]. Another included only SRC without comparison with non-SRC cancers[19]. The only study of its kind, the Taghavi *et al* study[10], did not find that SRC histology is an independent prognostic factor for death. The reason for this difference is not entirely clear. However, these two SEER analyses used data form different timeframe. The current study used SEER records from 2004 to 2020, whereas Taghavi *et al* used data from 2004 to 2007. The current report included 7,979 SRC (20.8%) and 30,357 non-SRC (79.2%) patients, compared with 2,666 (26.0%) SRC and 7,580 (74.0%) non-SRC in Taghavi’s analysis. The large sample size enables robust stratified and interaction analyses. ICD code 8490, which is an international standard, was both used for defining SRC patients in these two analyses. We also found, more than 97% SRC are poorly differentiated gastric cancer in SEER from 2004 to 2020, indicating consistent diagnosis of SRC. Taghavi *et al* only used the code ICD 8140 as inclusion criteria for non-SRC gastric adenocarcinoma. We additionally included ICD code 8144/3, 8145/3, 8211/3, 8260/3, 8480/3 for non-SRC adenocarcinomas. Thus, our sample size and proportion of non-SRC patients were larger.

Our findings have some limitations. First, this is a retrospective study based on registration databases, thus bias is unavoidable. The SEER is a large, population-based cohort which is representative of the US population, thus reduce the effect of selection bias. Previous studies reported conflicting results, at least partially due to numerous challenges exist in unveiling the prognostication of SRC histology. For example, the rarity of this disease, heterogeneous study population and different extent of disease at presentation in different studies[10]. The large population-based SEER database can be an optimal source to circumvent these challenges. The updated large sample size in the current report allows comprehensive statistical analyses such as interaction analyses and matching for important prognostic determinators.

In conclusion, this population-based study reveals that SRC histology is an independent predictor of poor prognosis in gastric adenocarcinoma. Although SRC significantly differ from non-SRC cancers in extent of disease at presentation, stage for stage, SRC still confers worse prognosis. SRC histology is a biologically aggressive entity of gastric cancer, however, early stage SRC might live longer because of significantly younger age of diagnosis.

## Data Availability

All data produced in the present study are available upon reasonable request to the authors
All data produced in the present work are contained in the manuscript

## References

1. Sung H, Ferlay J, Siegel RL, et al. Global cancer statistics 2020: Globocan estimates of incidence and mortality worldwide for 36 cancers in 185 countries. CA Cancer J Clin. 2021; 71: 209–249.

2. Van Cutsem E, Sagaert X, Topal B, et al. Gastric cancer. Lancet. 2016; 388: 2654–2664.

3. Benesch MGK, Mathieson A. Epidemiology of signet ring cell adenocarcinomas. Cancers (Basel). 2020; 12:

4. Thrift AP, El-Serag HB. Burden of gastric cancer. Clinical Gastroenterology and Hepatology. 2020; 18: 534–542.

5. Wong MCS, Huang J, Chan PSF, et al. Global incidence and mortality of gastric cancer, 1980-2018. JAMA Netw Open. 2021; 4: e2118457.

6. H watanabe, jr jass, lh sobin, etal : Histological typing of oesophageal and gastric tumours: Who international histological classification of tumours no. 18 1990 ed 2 berlin, germany springer. 1990;

7. Lauren P. The two histological main types of gastric carcinoma: Diffuse and so-called intestinal-type carcinoma. An attempt at a histo-clinical classification. Acta Pathol Microbiol Scand. 1965; 64: 31–49.

8. Ribeiro MM, Sarmento JA, Sobrinho Simões MA, et al. Prognostic significance of lauren and ming classifications and other pathologic parameters in gastric carcinoma. Cancer. 1981; 47: 780–4.

9. Piessen G, Messager M, Leteurtre E, et al. Signet ring cell histology is an independent predictor of poor prognosis in gastric adenocarcinoma regardless of tumoral clinical presentation. Ann Surg. 2009; 250: 878–87.

10. Taghavi S, Jayarajan SN, Davey A, et al. Prognostic significance of signet ring gastric cancer. J Clin Oncol. 2012; 30: 3493–8.

11. Chon HJ, Hyung WJ, Kim C, et al. Differential prognostic implications of gastric signet ring cell carcinoma: Stage adjusted analysis from a single high-volume center in asia. Ann Surg. 2017; 265: 946–953.

12. Gronnier C, Messager M, Robb WB, et al. Is the negative prognostic impact of signet ring cell histology maintained in early gastric adenocarcinoma? Surgery. 2013; 154: 1093–9.

13. Kim HW, Kim JH, Lim BJ, et al. Sex disparity in gastric cancer: Female sex is a poor prognostic factor for advanced gastric cancer. Ann Surg Oncol. 2016; 23: 4344–4351.

14. Ma J, Meng Y, Zhou X, et al. The prognostic significance and gene expression characteristics of gastric signet-ring cell carcinoma: A study based on the seer and tcga databases. Front Surg. 2022; 9: 819018.

15. Graziosi L, Marino E, Natalizi N, et al. Prognostic survival significance of signet ring cell (src) gastric cancer: Retrospective analysis from a single western center. J Pers Med. 2023; 13:

16. Huang KH, Chen MH, Fang WL, et al. The clinicopathological characteristics and genetic alterations of signet-ring cell carcinoma in gastric cancer. Cancers (Basel). 2020; 12:

17. Ha TK, An JY, Youn HK, et al. Indication for endoscopic mucosal resection in early signet ring cell gastric cancer. Ann Surg Oncol. 2008; 15: 508–13.

18. Curtis NJ, Noble F, Bailey IS, et al. The relevance of the siewert classification in the era of multimodal therapy for adenocarcinoma of the gastro-oesophageal junction. J Surg Oncol. 2014; 109: 202–7.

19. Wei Q, Gao Y, Qi C, et al. Clinicopathological characteristics and prognosis of signet ring gastric cancer: A population-based study. Front Oncol. 2021; 11: 580545.

20. Jiang CG, Wang ZN, Sun Z, et al. Clinicopathologic characteristics and prognosis of signet ring cell carcinoma of the stomach: Results from a chinese mono-institutional study. J Surg Oncol. 2011; 103: 700–3.

21. Kunisaki C, Shimada H, Nomura M, et al. Therapeutic strategy for signet ring cell carcinoma of the stomach. Br J Surg. 2004; 91: 1319–24.

22. Kwon KJ, Shim KN, Song EM, et al. Clinicopathological characteristics and prognosis of signet ring cell carcinoma of the stomach. Gastric Cancer. 2014; 17: 43–53.

23. Kim HM, Pak KH, Chung MJ, et al. Early gastric cancer of signet ring cell carcinoma is more amenable to endoscopic treatment than is early gastric cancer of poorly differentiated tubular adenocarcinoma in select tumor conditions. Surg Endosc. 2011; 25: 3087–93.

24. Zhao S, Lv L, Zheng K, et al. Prognosis and biological behavior of gastric signet-ring cell carcinoma better or worse: A meta-analysis. Front Oncol. 2021; 11: 603070.

25. Zaafouri H, Jouini R, Khedhiri N, et al. Comparison between signet-ring cell carcinoma and non-signet-ring cell carcinoma of the stomach: Clinicopathological parameters, epidemiological data, outcome, and prognosis-a cohort study of 123 patients from a non-endemic country. World J Surg Oncol. 2022; 20: 238.

26. Institute NC. Surveillance, epidemiology, and end results (seer) program. Cancer Statistics, SEER Data & Software, Registry Operations. 2018;

27. Drubay V, Nuytens F, Renaud F, et al. Poorly cohesive cells gastric carcinoma including signet-ring cell cancer: Updated review of definition, classification and therapeutic management. World J Gastrointest Oncol. 2022; 14: 1406–1428.

28. Oota K. Histological typing of gastric and oesophageal tumors. Inter national Histological Classification of Tumours No18. 1977; 19:

29. Bosman FT, Carneiro F, Hruban RH, et al. Who classification of tumours of the digestive system. Journal. 2010;

30. Nagtegaal ID, Odze RD, Klimstra D, et al. The 2019 who classification of tumours of the digestive system. Histopathology. 2020; 76: 182.

31. Ming SC. Gastric carcinoma: A pathobiological classification. Cancer. 1977; 39: 2475–2485.

32. Chen J, Liu K, Luo Y, et al. Single-cell profiling of tumor immune microenvironment reveals immune irresponsiveness in gastric signet-ring cell carcinoma. Gastroenterology. 2023; 165: 88–103.

33. Puccini A, Poorman K, Catalano F, et al. Molecular profiling of signet-ring-cell carcinoma (srcc) from the stomach and colon reveals potential new therapeutic targets. Oncogene. 2022; 41: 3455–3460.

34. Zhao W, Jia Y, Sun G, et al. Single-cell analysis of gastric signet ring cell carcinoma reveals cytological and immune microenvironment features. Nat Commun. 2023; 14: 2985.

35. Kunz PL, Gubens M, Fisher GA, et al. Long-term survivors of gastric cancer: A california population-based study. J Clin Oncol. 2012; 30: 3507–15.

