## Supplemental Table 1-11 for "Signet ring cell histology is an independent predictor of poor prognosis in gastric adenocarcinoma: A population-based analysis"

Table S1. Association between clinical characteristics with survival for patients with gastric adenocarcinoma after propensity score matching[c], SEER 2004-2020.

| Characteristic | N | mOS | Overall Survival: Univariable HR | 95% CI | *P* | Overall Survival: Multivariable[a] HR | 95% CI | *P* | Cause-specific Survival: Univariable HR | 95% CI | *P* | Cause-specific Survival: Multivariable[a] HR | 95% CI | *P* |
|---|---|---|---|---|---|---|---|---|---|---|---|---|---|---|
| Age | 15832 | 16 | 1.01 | 1.0-1.01 | < 0.001 | | | | 1.0 | 1.0-1.0 | 0.274 | | | |
| Age, y | | | | | | | | | | | | | | |
| < 65 (ref.) | 9615 | 17 | 1.0 | 1.0 | | 1.0 | 1.0 | | 1.0 | 1.0 | | 1.0 | 1.0 | |
| >= 65 | 6217 | 15 | 1.19 | 1.15-1.23 | < 0.001 | 1.29 | 1.24-1.34 | < 0.001 | 1.05 | 1.01-1.09 | 0.009 | 1.16 | 1.12-1.21 | < 0.001 |
| Histology | | | | | | | | | | | | | | |
| Gastric non-SRC (ref.) | 7916 | 18 | 1.0 | 1.0 | | 1.0 | 1.0 | | 1.0 | 1.0 | | 1.0 | 1.0 | |
| Gastric SRC | 7916 | 15 | 1.12 | 1.08-1.16 | < 0.001 | 1.18 | 1.14-1.23 | < 0.001 | 1.16 | 1.12-1.21 | < 0.001 | 1.23 | 1.18-1.27 | < 0.001 |
| Sex | | | | | | | | | | | | | | |
| Male (ref.) | 8048 | 16 | 1.0 | 1.0 | | / | / | | 1.0 | 1.0 | | / | / | |
| Female | 7784 | 16 | 1 | 0.96-1.03 | 0.826 | / | / | / | 1.0 | 0.97-1.04 | 0.867 | / | / | / |
| Race/ethnicity | | | | | | | | | | | | | | |
| White | 6729 | 16 | 1.0 | 1.0 | | 1.0 | 1.0 | | 1.0 | 1.0 | | 1.0 | 1.0 | |
| Black | 1691 | 17 | 0.93 | 0.88-0.99 | 0.028 | 1.04 | 0.98-1.11 | 0.173 | 0.92 | 0.86-0.98 | 0.014 | 1.01 | 0.95-1.08 | 0.718 |
| API | 2840 | 22 | 0.74 | 0.71-0.78 | < 0.001 | 0.85 | 0.8-0.89 | < 0.001 | 0.77 | 0.73-0.81 | < 0.001 | 0.87 | 0.82-0.92 | < 0.001 |
| AIA | 141 | 14 | 0.98 | 0.81-1.19 | 0.822 | 1.15 | 0.95-1.39 | 0.156 | 1 | 0.82-1.23 | 0.972 | 1.17 | 0.95-1.43 | 0.138 |
| Hispanic | 4431 | 15 | 0.97 | 0.93-1.01 | 0.189 | 0.95 | 0.91-0.99 | 0.023 | 1.02 | 0.97-1.07 | 0.412 | 0.96 | 0.92-1.01 | 0.126 |
| Primary site | | | | | | | | | | | | | | |
| Upper stomach | 3269 | 15 | 1.0 | 1.0 | | 1.0 | 1.0 | | 1.0 | 1.0 | | 1.0 | 1.0 | |
| Middle stomach | 4390 | 20 | 0.75 | 0.71-0.79 | < 0.001 | 0.93 | 0.88-0.98 | 0.011 | 0.75 | 0.71-0.79 | < 0.001 | 0.93 | 0.88-0.99 | 0.015 |
| Lower stomach | 4094 | 22 | 0.75 | 0.71-0.79 | < 0.001 | 0.96 | 0.91-1.02 | 0.155 | 0.73 | 0.69-0.77 | < 0.001 | 0.95 | 0.9-1.01 | 0.102 |
| Overlapping | 4079 | 12 | 1.19 | 1.13-1.25 | < 0.001 | 1.11 | 1.06-1.18 | < 0.001 | 1.2 | 1.13-1.26 | < 0.001 | 1.1 | 1.04-1.16 | 0.001 |
| Stage | | | | | | | | | | | | | | |
| Ⅰ | 3720 | 87 | 1 | 1 | | 1.0 | 1.0 | | 1.0 | 1.0 | | 1.0 | 1.0 | |
| Ⅱ | 2195 | 36 | 1.48 | 1.38-1.59 | < 0.001 | 1.95 | 1.81-2.1 | < 0.001 | 1.78 | 1.65-1.93 | < 0.001 | 2.33 | 2.14-2.53 | < 0.001 |
| Ⅲ | 2341 | 21 | 2.23 | 2.09-2.38 | < 0.001 | 3.08 | 2.88-3.3 | < 0.001 | 2.81 | 2.61-3.02 | < 0.001 | 3.85 | 3.56-4.16 | < 0.001 |
| Ⅳ | 7576 | 9 | 4.98 | 4.72-5.25 | < 0.001 | 4.37 | 4.11-4.64 | < 0.001 | 6.51 | 6.12-6.92 | < 0.001 | 5.52 | 5.16-5.91 | < 0.001 |

| | | | | | | | | | | | | | | |
|---|---|---|---|---|---|---|---|---|---|---|---|---|---|---|
| *P*-trend | | | 1.72 | 1.69-1.75 | < 0.001 | 1.60 | 1.57-1.63 | < 0.001 | 1.87 | 1.84-1.91 | < 0.001 | 1.70 | 1.66-1.73 | < 0.001 |
| Peritoneal metastasis[b] | | | | | | | | | | | | | | |
| Negative | 8035 | 26 | 1.0 | 1.0 | | | | | 1.0 | 1.0 | | | | |
| Positive | 3710 | 7 | 3.29 | 3.15-3.43 | < 0.001 | | | | 3.59 | 3.43-3.76 | < 0.001 | | | |
| Table S1. Continued | | | | | | | | | | | | | | |
| Surgery | | | | | | | | | | | | | | |
| No (ref.) | 6722 | 8 | 1.0 | 1.0 | | 1.0 | 1.0 | | 1.0 | 1.0 | | 1.0 | 1.0 | |
| Yes | 9110 | 34 | 0.27 | 0.26-0.28 | < 0.001 | 0.37 | 0.35-0.39 | < 0.001 | 0.25 | 0.24-0.26 | < 0.001 | 0.36 | 0.35-0.38 | < 0.001 |
| Radiation therapy | | | | | | | | | | | | | | |
| No (ref.) | 11463 | 14 | 1.0 | 1.0 | | 1.0 | 1.0 | | 1.0 | 1.0 | | 1.0 | 1.0 | |
| Yes | 4369 | 22 | 0.77 | 0.74-0.81 | < 0.001 | 1.06 | 1.01-1.11 | 0.051 | 0.77 | 0.73-0.8 | < 0.001 | 1.06 | 1.01-1.11 | 0.019 |
| Chemotherapy | | | | | | | | | | | | | | |
| No (ref.) | 4794 | 16 | 1.0 | 1.0 | | 1.0 | 1.0 | | 1.0 | 1.0 | | 1.0 | 1.0 | |
| Yes | 11038 | 16 | 1.11 | 1.07-1.16 | < 0.001 | 0.6 | 0.57-0.63 | < 0.001 | 1.22 | 1.17-1.28 | < 0.001 | 0.61 | 0.58-0.64 | < 0.001 |

Abbreviations: SEER: Surveillance, Epidemiology, End Results; SRC: signet ring cell carcinoma; API: Asian/Pacific Islander; AIA: American Indian/Alaska Native

[a] Variables included in the propensity score were age(>= 65, < 65 years old), sex, race, primary site, stage, surgery, radiation therapy and chemotherapy.

[b] Data for peritoneal metastases were used from 2004 to 2015 because data after 2015 could not separate out peritoneal cytological positive groups.

Table S2. Association between clinical characteristics with mortality for patients with gastric adenocarcinoma with peritoneal cytology in multivariable analysis, SEER 2004-2015[a].

| Clinical Characteristics | N | mOS | Overall Survival | | | | | | Cancer-specific Survival | | | | | |
|---|---|---|---|---|---|---|---|---|---|---|---|---|---|---|
| | | | Univariable | | | Multivariable[b] | | | Univariable | | | Multivariable[c] | | |
| | | | HR | 95% CI | *P* | HR | 95% CI | *P* | HR | 95% CI | *P* | HR | 95% CI | *P* |
| Age | 27672 | 18 | 1.01 | 1.01-1.01 | < 0.001 | / | / | / | 1.00 | 1.00-1.00 | 0.717 | / | / | / |
| Age, y | | | | | | | | | | | | | | |
| <65 | 12634 | 18 | 1.0 | 1.0 | | 1.0 | 1.0 | | 1.0 | 1.0 | | | / | |
| >=65 | 15038 | 17 | 1.17 | 1.14-1.2 | < 0.001 | 1.3 | 1.26-1.34 | < 0.001 | 0.99 | 0.96-1.02 | 0.355 | / | / | / |
| Histology | | | | | | | | | | | | | | |
| Gastric non-SRC (ref.) | 21734 | 19 | 1.0 | 1.0 | | 1.0 | 1.0 | | 1.0 | 1.0 | | 1.0 | 1.0 | |
| Gastric SRC | 5938 | 15 | 1.13 | 1.1-1.17 | < 0.001 | 1.2 | 1.16-1.24 | < 0.001 | 1.24 | 1.19-1.28 | < 0.001 | 1.24 | 1.2-1.28 | < 0.001 |
| Sex | | | | | | | | | | | | | | |
| Male (ref.) | 17777 | 18 | 1.0 | 1.0 | | / | / | | 1.0 | 1.0 | | / | / | |
| Female | 9895 | 17 | 0.99 | 0.96-1.02 | 0.396 | / | / | / | 0.99 | 0.96-1.02 | 0.382 | / | / | / |
| Race/ethnicity | | | | | | | | | | | | | | |
| White | 14001 | 17 | 1.0 | 1.0 | | 1.0 | 1.0 | | 1.0 | 1.0 | | 1.0 | 1.0 | |
| Black | 3355 | 16 | 0.97 | 0.93-1.01 | 0.17 | 1.03 | 0.99-1.08 | 0.147 | 0.96 | 0.91-1 | 0.06 | 1 | 0.95-1.04 | 0.836 |
| API | 4625 | 26 | 0.71 | 0.68-0.74 | < 0.001 | 0.79 | 0.76-0.82 | < 0.001 | 0.72 | 0.69-0.75 | < 0.001 | 0.82 | 0.78-0.86 | < 0.001 |
| AIA | 250 | 14 | 1.09 | 0.95-1.25 | 0.209 | 1.18 | 1.03-1.35 | 0.019 | 1.14 | 0.98-1.32 | 0.081 | 1.18 | 1.02-1.36 | 0.03 |
| Hispanic | 5441 | 17 | 0.9 | 0.87-0.94 | < 0.001 | 0.91 | 0.87-0.94 | < 0.001 | 0.95 | 0.92-0.99 | 0.012 | 0.92 | 0.88-0.95 | < 0.001 |
| Primary site | | | | | | | | | | | | | | |
| Upper stomach | 10299 | 17 | 1.0 | 1.0 | | 1.0 | 1.0 | | 1.0 | 1.0 | | 1.0 | 1.0 | |
| Middle stomach | 5956 | 22 | 0.81 | 0.78-0.84 | < 0.001 | 0.97 | 0.93-1.01 | 0.111 | 0.78 | 0.75-0.81 | < 0.001 | 0.96 | 0.92-1.01 | 0.089 |
| Lower stomach | 6620 | 24 | 0.79 | 0.76-0.82 | < 0.001 | 1.02 | 0.98-1.06 | 0.296 | 0.75 | 0.72-0.78 | < 0.001 | 1.02 | 0.98-1.07 | 0.285 |
| Overlapping | 4797 | 12 | 1.24 | 1.19-1.28 | < 0.001 | 1.22 | 1.17-1.27 | < 0.001 | 1.26 | 1.21-1.31 | < 0.001 | 1.22 | 1.17-1.28 | < 0.001 |
| Peritoneal cytology | | | | | | | | | | | | | | |
| Negative | 20175 | 28 | 1.0 | 1.0 | | 1.0 | 1.0 | | 1.0 | 1.0 | | 1.0 | 1.0 | |
| Positive | 7497 | 7 | 3.43 | 3.33-3.53 | < 0.001 | 2.14 | 2.06-2.22 | < 0.001 | 3.87 | 3.75-3.99 | < 0.001 | 2.21 | 2.13-2.3 | < 0.001 |

| | | | | | | | | | | | | | | |
|---|---|---|---|---|---|---|---|---|---|---|---|---|---|---|
| Surgery | | | | | | | | | | | | | | |
| No (ref.) | 11050 | 8 | 1.0 | 1.0 | | 1.0 | 1.0 | | 1.0 | 1.0 | | 1.0 | 1.0 | |
| Yes | 16622 | 38 | 0.27 | 0.27-0.28 | < 0.001 | 0.36 | 0.35-0.37 | < 0.001 | 0.25 | 0.24-0.25 | < 0.001 | 0.34 | 0.33-0.35 | < 0.001 |
| Radiation therapy | | | | | | | | | | | | | | |
| No (ref.) | 18696 | 15 | 1.0 | 1.0 | | 1.0 | 1.0 | | 1.0 | 1.0 | | 1.0 | 1.0 | |
| Yes | 8976 | 22 | 0.85 | 0.82-0.87 | < 0.001 | 1.1 | 1.07-1.14 | 0.176 | 0.87 | 0.84-0.9 | < 0.001 | 1.13 | 1.09-1.17 | 0.061 |
| Chemotherapy | | | | | | | | | | | | | | |
| No (ref.) | 10988 | 19 | 1.0 | 1.0 | | 1.0 | 1.0 | | 1.0 | 1.0 | | 1.0 | 1.0 | |
| Yes | 16684 | 17 | 1.15 | 1.12-1.18 | < 0.001 | 0.82 | 0.79-0.85 | < 0.001 | 1.33 | 1.29-1.38 | < 0.001 | 0.85 | 0.82-0.88 | < 0.001 |

Abbreviation: SEER: Surveillance, Epidemiology, End Results; ref.: referent; HR: hazards ratio; CI: Confidence interval; mOS: median overall survival; SRC: signet ring cell carcinoma; API: Asian/Pacific Islander; AIA: American Indian/Alaska Native

[a] Data were used because data after 2015 did not include information of peritoneal cytology.

[b] Variables included in multivariate analysis for overall survival were age (>= 65, < 65 years old), primary site, histology, race, peritoneal cytology, surgery, radiation therapy and chemotherapy.

[c] Variables included in multivariate analysis for cause-specific survival were primary site, histology, race, stage, surgery, radiation therapy and chemotherapy.

Table S3. Baseline characteristics of gastric cancer (signet ring cell carcinoma and non-signet ring cell carcinoma) groups after propensity score matching[a] with peritoneal cytology, SEER 2004-2015[b]

| Clinical Characteristics | All Gastric cancer (N=11672, %) | SRC (N=5836, %) | non-SRC (N=5836, %) | $P^c$ |
|---|---|---|---|---|
| OS | | | | |
| Alive | 2276(19.5) | 1058(18.1) | 1218(20.9) | < 0.001 |
| Dead | 9396(80.5) | 4778(81.9) | 4618(79.1) | < 0.001 |
| CSS | | | | |
| Alive or dead of other cause | 3385(29) | 1518(26.0) | 1867(32.0) | < 0.001 |
| Dead (attributable to this cancer dx) | 8287(71) | 4318(74.0) | 3969(68.0) | < 0.001 |
| Survival months, Mean (SD) | 38.86(48.13) | 36.01(46.64) | 41.70(49.41) | < 0.001 |
| Age, y | | | | |
| Mean (SD) | 61.04(13.98) | 60.09(14.15) | 62.00(13.75) | <0.001 |
| <65 | 6988(59.9) | 3500(60.0) | 3488(59.8) | |
| >=65 | 4684(40.1) | 2336(40.0) | 2348(40.2) | 0.835 |
| Sex | | | | 0.97 |
| Male | 6135(52.6) | 3066(52.5) | 3069(52.6) | |
| Female | 5537(47.4) | 2770(47.5) | 2767(47.4) | |
| Race/ethnicity | | | | |
| White | 5145(44.1) | 2562(43.9) | 2583(44.3) | 0.709 |
| Black | 1351(11.6) | 682(11.7) | 669(11.5) | 0.728 |
| API | 2040(17.5) | 1010(17.3) | 1030(17.6) | 0.643 |
| AIA | 97(0.8) | 51(0.9) | 46(0.8) | 0.683 |
| Hispanic | 3039(26) | 1531(26.2) | 1508(25.8) | 0.643 |
| Primary site | | | | |
| Upper stomach | 2513(21.5) | 1258(21.6) | 1255(21.5) | 0.964 |
| Middle stomach | 3239(27.8) | 1642(28.1) | 1597(27.4) | 0.363 |
| Lower stomach | 3058(26.2) | 1521(26.1) | 1537(26.3) | 0.752 |
| Overlapping | 2862(24.5) | 1415(24.2) | 1447(24.8) | 0.505 |
| Peritoneal cytology[c] | | | | |
| Negative | 8139(69.7) | 4035(69.1) | 4104(70.3) | |
| Positive | 3533(30.3) | 1801(30.9) | 1732(29.7) | 0.171 |
| Surgery | | | | |
| No | 4675(40.1) | 2370(40.6) | 2305(39.5) | |
| Yes | 6997(59.9) | 3466(59.4) | 3531(60.5) | 0.227 |
| Radiation therapy | | | | |
| No | 8155(69.9) | 4075(69.8) | 4080(69.9) | |
| Yes | 3517(30.1) | 1761(30.2) | 1756(30.1) | 0.936 |
| Chemotherapy | | | | |
| No | 3842(32.9) | 1931(33.1) | 1911(32.7) | |
| Yes | 7830(67.1) | 3905(66.9) | 3925(67.3) | 0.708 |

Abbreviation: SEER: Surveillance, Epidemiology, End Results; SRC: signet ring cell carcinoma; API: Asian/Pacific Islander; AIA: American Indian/Alaska Native

[a] Variables included in the propensity score were age ((>= 65, < 65 years old), sex, race, primary site, peritoneal cytology, surgery, radiation therapy and chemotherapy.

[b] Data from 2004 to 2015 for peritoneal metastases were used because data after 2015 did not include information of peritoneal cytology.

[c] Derived from $\chi2$-test for categorical variables, general linear models for continuous variables.

Table S4.1. Heterogeneity in the association of survival with clinical characteristics between patients with gastric signet ring cell carcinoma and non-signet ring cell carcinoma after propensity score matching[a], SEER 2004-2020

| Clinical characristics | N, non-SRC (ref.) | N, SRC | OS | | | | | CSS | | | | |
|---|---|---|---|---|---|---|---|---|---|---|---|---|
| | | | mOS (non-SRC) | mOS (SRC) | HR | 95% CI | *P*-*heterogeneity* | mCSS (non-SRC) | mCSS (SRC) | HR | 95% CI | *P*-*heterogeneity* |
| Age (as continuous variable) | 7978 | 7978 | 17 | 15 | | | **0.078** | 18 | 16 | | | **0.001** |
| Sex | | | | | | | 0.323 | | | | | 0.269 |
| Male | 5130 | 4017 | 18 | 15 | 1.13 | 1.08-1.18 | | 20 | 15 | 1.17 | 1.11-1.23 | |
| Female | 2846 | 3959 | 17 | 15 | 1.09 | 1.03-1.15 | | 19 | 16 | 1.12 | 1.06-1.19 | |
| Race/ethnicity | | | | | | | 0.056 | | | | | 0.06 |
| White | 3076 | 3388 | 17 | 14 | 1.1 | 1.04-1.16 | | 18 | 15 | 1.13 | 1.06-1.19 | |
| Black | 872 | 841 | 18 | 15 | 1.12 | 1.01-1.25 | | 20 | 17 | 1.16 | 1.03-1.3 | |
| API | 2082 | 1415 | 24 | 20 | 1.06 | 0.97-1.15 | | 27 | 21 | 1.09 | 1-1.19 | |
| AIA | 112 | 76 | 12 | 15 | 0.95 | 0.68-1.33 | | 13 | 15 | 0.99 | 0.7-1.41 | |
| Hispanic | 1814 | 2236 | 17 | 13 | 1.22 | 1.14-1.32 | | 18 | 14 | 1.27 | 1.18-1.37 | |
| Primary site | | | | | | | **0.008** | | | | | **0.044** |
| Upper stomach | 3002 | 1634 | 18 | 13 | 1.26 | 1.17-1.34 | | 19 | 14 | 1.29 | 1.2-1.38 | |
| Middle stomach | 1723 | 2212 | 22 | 18 | 1.09 | 1.01-1.17 | | 24 | 19 | 1.13 | 1.04-1.22 | |
| Lower stomach | 1850 | 2050 | 23 | 20 | 1.08 | 1-1.16 | | 27 | 22 | 1.15 | 1.06-1.24 | |
| Overlapping | 1404 | 2083 | 12 | 12 | 1.13 | 1.05-1.22 | | 13 | 12 | 1.16 | 1.07-1.25 | |
| Stage | | | | | | | **0.076** | | | | | 0.335 |
| Ⅰ | 2185 | 1866 | 89 | 89 | 1.08 | 0.99-1.18 | | | | 1.29 | 1.16-1.44 | |
| Ⅱ | 1230 | 1101 | 38 | 33 | 1.15 | 1.04-1.28 | | 45 | 36 | 1.21 | 1.08-1.35 | |
| Ⅲ | 1024 | 1202 | 23 | 19 | 1.26 | 1.14-1.38 | | 26 | 20 | 1.28 | 1.15-1.41 | |
| Ⅳ | 3524 | 3794 | 10 | 8 | 1.14 | 1.09-1.2 | | 10 | 8 | 1.15 | 1.1-1.21 | |
| Peritoneal cytology[b] | | | | | | | 0.089 | | | | | 0.947 |
| Negative | 3977 | 4058 | 29 | 23 | 1.11 | 1.05-1.17 | | 35 | 26 | 1.18 | 1.12-1.25 | |
| Positive | 1861 | 1849 | 8 | 7 | 1.18 | 1.11-1.26 | | 8 | 7 | 1.19 | 1.11-1.27 | |
| Surgery | | | | | | | 0.877 | | | | | 0.132 |
| No | 3360 | 3360 | 8 | 8 | 1.13 | 1.07-1.19 | | 9 | 8 | 1.14 | 1.09-1.2 | |
| Yes | 4565 | 4565 | 41 | 31 | 1.15 | 1.09-1.21 | | 52 | 35 | 1.22 | 1.16-1.3 | |
| Radiation therapy | | | | | | | 0.371 | | | | | 0.569 |
| No | 5686 | 5756 | 16 | 13 | 1.1 | 1.06-1.15 | | 17 | 14 | 1.14 | 1.09-1.19 | |
| Yes | 2239 | 2169 | 23 | 20 | 1.15 | 1.08-1.24 | | 27 | 21 | 1.18 | 1.1-1.27 | |
| Chemotherapy | | | | | | | **< 0.001** | | | | | **0.015** |
| No | 2649 | 2412 | 18 | 15 | 1 | 0.94-1.06 | | 22 | 18 | 1.06 | 0.99-1.14 | |
| Yes | 5276 | 5513 | 18 | 15 | 1.19 | 1.13-1.24 | | 19 | 15 | 1.2 | 1.15-1.26 | |

Abbreviations: SEER: Surveillance, Epidemiology, End Results; ref.: referent; HR: hazards ratio; CI: Confidence interval; mOS: median overall survival; mCSS: median cause-specific survival; SRC: signet ring cell carcinoma; API: Asian/Pacific Islander; AIA: American Indian/Alaska Native

[a] Adjusted for propensity score which included in for the propensity score were age (>= 65, < 65years old), sex, race, primary site, stage, surgery, radiation therapy and chemotherapy, however, the same variable were excluded from the score for the analyses of specific clinical characteristics.

[b] Data from 2004 to 2015 for peritoneal metastases were used because data after 2015 did not include information of peritoneal cytology.

Table S4.2. Heterogeneity in the association of survival with clinical characteristics between patients with gastric signet ring cell carcinoma and non-signet ring cell carcinoma after propensity score matching[a], SEER 2004-2020.

| Clinical characristics | N, non-SRC (ref.) | N, SRC | OS | | | | | CSS | | | | |
|---|---|---|---|---|---|---|---|---|---|---|---|---|
| | | | mOS (non-SRC) | mOS (SRC) | HR[b] | 95% CI | *P*-*heterogeneity*[b] | mCSS (non-SRC) | mCSS (SRC) | HR[b] | 95% CI | *P*-*heterogeneity*[b] |
| Age (as continuous variable) | 7916 | 7916 | 18 | 15 | | | 0.581 | 20 | 16 | | | 0.658 |
| Sex | | | | | | | 0.104 | | | | | 0.054 |
| Male | 4030 | 4018 | 19 | 15 | 1.24 | 1.18-1.3 | | 20 | 15 | 1.28 | 1.21-1.35 | |
| Female | 3886 | 3898 | 18 | 15 | 1.17 | 1.11-1.23 | | 19 | 16 | 1.19 | 1.13-1.26 | |
| Race/ethnicity | | | | | | | 0.676 | | | | | 0.672 |
| White | 3366 | 3363 | 17 | 14 | 1.23 | 1.16-1.29 | | 19 | 15 | 1.25 | 1.18-1.33 | |
| Black | 844 | 847 | 19 | 15 | 1.23 | 1.1-1.37 | | 20 | 17 | 1.26 | 1.12-1.42 | |
| API | 1423 | 1417 | 24 | 20 | 1.16 | 1.06-1.27 | | 27 | 21 | 1.18 | 1.07-1.29 | |
| AIA | 65 | 76 | 12 | 15 | 1.26 | 0.84-1.89 | | 12 | 15 | 1.29 | 0.84-1.99 | |
| Hispanic | 2218 | 2213 | 16 | 13 | 1.18 | 1.1-1.27 | | 17 | 14 | 1.23 | 1.14-1.32 | |
| Primary site | | | | | | | 0.881 | | | | | 0.925 |
| Upper stomach | 1635 | 1634 | 17 | 13 | 1.23 | 1.14-1.33 | | 18 | 14 | 1.26 | 1.16-1.37 | |
| Middle stomach | 2199 | 2191 | 22 | 18 | 1.2 | 1.11-1.29 | | 25 | 19 | 1.22 | 1.13-1.32 | |
| Lower stomach | 2053 | 2041 | 24 | 20 | 1.2 | 1.12-1.3 | | 28 | 22 | 1.25 | 1.16-1.35 | |
| Overlapping | 2029 | 2050 | 13 | 11 | 1.2 | 1.12-1.29 | | 13 | 12 | 1.23 | 1.14-1.32 | |
| Stage | | | | | | | **0.022** | | | | | 0.703 |
| Ⅰ | 1850 | 1870 | 83 | 90 | 1.16 | 1.06-1.27 | | | | 1.29 | 1.15-1.44 | |
| Ⅱ | 1092 | 1103 | 41 | 33 | 1.29 | 1.16-1.43 | | 51 | 36 | 1.32 | 1.18-1.49 | |
| Ⅲ | 1177 | 1164 | 23 | 18 | 1.29 | 1.17-1.41 | | 26 | 20 | 1.3 | 1.18-1.44 | |
| Ⅳ | 3797 | 3779 | 10 | 8 | 1.18 | 1.13-1.24 | | 10 | 8 | 1.19 | 1.14-1.25 | |
| Peritoneal cytology[c] | | | | | | | 0.205 | | | | | 0.823 |
| Negative | 3977 | 4058 | 29 | 23 | 1.13 | 1.07-1.19 | | 35 | 26 | 1.19 | 1.13-1.26 | |
| Positive | 1861 | 1849 | 8 | 7 | 1.16 | 1.09-1.24 | | 8 | 7 | 1.17 | 1.09-1.25 | |
| Surgery | | | | | | | 0.304 | | | | | 0.979 |
| No | 3344 | 3378 | 9 | 8 | 1.19 | 1.13-1.25 | | 9 | 8 | 1.21 | 1.15-1.27 | |
| Yes | 4572 | 4538 | 38 | 31 | 1.22 | 1.16-1.29 | | 49 | 35 | 1.27 | 1.2-1.34 | |
| Radiation therapy | | | | | | | 0.397 | | | | | 0.574 |
| No | 5725 | 5738 | 16 | 13 | 1.19 | 1.14-1.24 | | 17 | 14 | 1.22 | 1.17-1.28 | |
| Yes | 2191 | 2178 | 24 | 20 | 1.24 | 1.16-1.33 | | 28 | 21 | 1.26 | 1.17-1.36 | |
| Chemotherapy | | | | | | | **< 0.001** | | | | | **0.013** |
| No | 2376 | 2418 | 18 | 15 | 1.09 | 1.02-1.17 | | 23 | 18 | 1.15 | 1.07-1.24 | |
| Yes | 5540 | 5498 | 18 | 15 | 1.25 | 1.2-1.31 | | 19 | 15 | 1.27 | 1.22-1.33 | |

Abbreviation: SEER: Surveillance, Epidemiology, End Results; ref.: referent; HR: hazards ratio; CI: Confidence interval; mOS: median overall survival; mCSS: median cause-specific survival; SRC: signet ring cell carcinoma; API: Asian/Pacific Islander; AIA: American Indian/Alaska Native

[a] Variables included in the propensity score were age (>= 65, < 65 years old), sex, race, primary site, stage, surgery, radiation therapy and chemotherapy.

[b] Adjusted for age (continuous variables), sex, race, primary site, stage, surgery, radiation therapy and chemotherapy

[c] Data from 2004 to 2015 for peritoneal metastases were used because data after 2015 did not include information of peritoneal cytology.

Table S4.3. Heterogeneity in the association of survival with clinical characteristics between patients with gastric signet ring cell carcinoma and non-signet ring cell carcinoma after propensity score matching[a], SEER 2004-2020.

| Clinical characristics | N, non-SRC (ref.) | N, SRC | OS | | | | | CSS | | | | |
|---|---|---|---|---|---|---|---|---|---|---|---|---|
| | | | mOS (non-SRC) | mOS (SRC) | HR[b] | 95% CI | $P_{-heterogeneity}$[b] | mCSS (non-SRC) | mCSS (SRC) | HR[b] | 95% CI | $P_{-heterogeneity}$[b] |
| Age (as continuous variable) | 7925 | 7925 | 18 | 15 | | | 0.41 | 20 | 16 | | | 0.766 |
| Sex | | | | | | | 0.119 | | | | | 0.06 |
| Male | 5130 | 4017 | 18 | 15 | 1.24 | 1.18-1.3 | | 20 | 15 | 1.28 | 1.22-1.34 | |
| Female | 2846 | 3959 | 17 | 15 | 1.18 | 1.11-1.24 | | 19 | 16 | 1.2 | 1.13-1.27 | |
| Race/ethnicity | | | | | | | 0.78 | | | | | 0.753 |
| White | 3076 | 3388 | 17 | 14 | 1.24 | 1.18-1.31 | | 18 | 15 | 1.28 | 1.2-1.35 | |
| Black | 872 | 841 | 18 | 15 | 1.27 | 1.14-1.42 | | 20 | 17 | 1.31 | 1.16-1.47 | |
| API | 2082 | 1415 | 24 | 20 | 1.19 | 1.1-1.3 | | 27 | 21 | 1.22 | 1.11-1.33 | |
| AIA | 112 | 76 | 12 | 15 | 1.2 | 0.85-1.7 | | 13 | 15 | 1.27 | 0.88-1.83 | |
| Hispanic | 1814 | 2236 | 17 | 13 | 1.2 | 1.12-1.3 | | 18 | 14 | 1.24 | 1.15-1.34 | |
| Primary site | | | | | | | 0.086 | | | | | 0.246 |
| Upper stomach | 3002 | 1634 | 18 | 13 | 1.31 | 1.22-1.4 | | 19 | 14 | 1.34 | 1.25-1.44 | |
| Middle stomach | 1723 | 2212 | 22 | 18 | 1.21 | 1.12-1.3 | | 24 | 19 | 1.24 | 1.14-1.34 | |
| Lower stomach | 1850 | 2050 | 23 | 20 | 1.18 | 1.09-1.27 | | 27 | 22 | 1.23 | 1.13-1.34 | |
| Overlapping | 1404 | 2083 | 12 | 12 | 1.24 | 1.15-1.34 | | 13 | 12 | 1.26 | 1.17-1.37 | |
| Stage | | | | | | | 0.102 | | | | | 0.456 |
| Ⅰ | 2185 | 1866 | 89 | 89 | 1.17 | 1.07-1.28 | | | | 1.3 | 1.17-1.45 | |
| Ⅱ | 1230 | 1101 | 38 | 33 | 1.21 | 1.09-1.33 | | 45 | 36 | 1.24 | 1.11-1.38 | |
| Ⅲ | 1024 | 1202 | 23 | 19 | 1.32 | 1.2-1.45 | | 26 | 20 | 1.32 | 1.19-1.46 | |
| Ⅳ | 3524 | 3794 | 10 | 8 | 1.18 | 1.12-1.24 | | 10 | 8 | 1.19 | 1.13-1.25 | |
| Surgery | | | | | | | 0.317 | | | | | 0.823 |
| No | 3360 | 3360 | 8 | 8 | 1.2 | 1.14-1.26 | | 9 | 8 | 1.21 | 1.15-1.28 | |
| Yes | 4565 | 4565 | 41 | 31 | 1.22 | 1.16-1.29 | | 52 | 35 | 1.27 | 1.2-1.35 | |
| Radiation therapy | | | | | | | 0.414 | | | | | 0.566 |
| No | 5686 | 5756 | 16 | 13 | 1.21 | 1.16-1.26 | | 17 | 14 | 1.24 | 1.19-1.3 | |
| Yes | 2239 | 2169 | 23 | 20 | 1.26 | 1.17-1.35 | | 27 | 21 | 1.29 | 1.2-1.38 | |
| Chemotherapy | | | | | | | **< 0.001** | | | | | **0.039** |
| No | 2649 | 2412 | 18 | 15 | 1.12 | 1.05-1.2 | | 22 | 18 | 1.19 | 1.1-1.28 | |
| Yes | 5276 | 5513 | 18 | 15 | 1.27 | 1.21-1.32 | | 19 | 15 | 1.28 | 1.23-1.34 | |

Abbreviation: SEER: Surveillance, Epidemiology, End Results; ref.: referent; HR: hazards ratio; CI: Confidence interval; mOS: median overall survival; mCSS: median cause-specific survival; SRC: signet ring cell carcinoma; API: Asian/Pacific Islander; AIA: American Indian/Alaska Native

[a] When we calculated each variable, we adjusted for propensity score (age, sex, race, primary site, stage, surgery, radiation therapy and chemotherapy, stripping out the calculating variable), and age was treated as categorical variables.

[b] Adjusted for age (>= 65, < 65 years old), sex, race, primary site, stage, surgery, radiation therapy and chemotherapy.

Table S4.4. Heterogeneity in the association of survival with clinical characteristics between patients with gastric signet ring cell carcinoma and non-signet ring cell carcinoma, SEER 2004-2020.

| Clinical characristics | N, non-SRC (ref.) | N, SRC | OS | | | | | CSS | | | | |
|---|---|---|---|---|---|---|---|---|---|---|---|---|
| | | | mOS (non-SRC) | mOS (SRC) | HR [a] | 95% CI | $P_{heterogeneity}$ [a] | mCSS (non-SRC) | mCSS (SRC) | HR [a] | 95% CI | $P_{heterogeneity}$ [a] |
| Age (as continuous variable) | 30357 | 7979 | 19 | 15 | | | **< 0.001** | 22 | 16 | | | **0.03** |
| Sex | | | | | | | 0.626 | | | | | 0.313 |
| Male | 20553 | 4018 | 19 | 15 | 1.25 | 1.2-1.3 | | 22 | 15 | 1.3 | 1.25-1.36 | |
| Female | 9804 | 3961 | 20 | 15 | 1.2 | 1.15-1.26 | | 23 | 16 | 1.24 | 1.18-1.3 | |
| Race/ethnicity | | | | | | | 0.549 | | | | | 0.767 |
| White | 15532 | 3393 | 18 | 14 | 1.22 | 1.17-1.27 | | 20 | 15 | 1.27 | 1.21-1.33 | |
| Black | 3660 | 853 | 18 | 15 | 1.25 | 1.14-1.36 | | 20 | 17 | 1.29 | 1.18-1.42 | |
| API | 5076 | 1417 | 30 | 20 | 1.26 | 1.17-1.36 | | 37 | 21 | 1.3 | 1.2-1.4 | |
| AIA | 270 | 76 | 13 | 15 | 1.1 | 0.82-1.48 | | 14 | 15 | 1.14 | 0.83-1.57 | |
| Hispanic | 5819 | 2240 | 19 | 13 | 1.2 | 1.13-1.28 | | 21 | 14 | 1.26 | 1.18-1.34 | |
| Primary site | | | | | | | 0.425 | | | | | 0.967 |
| Upper stomach | 12731 | 1634 | 18 | 13 | 1.27 | 1.2-1.35 | | 20 | 14 | 1.31 | 1.23-1.39 | |
| Middle stomach | 6071 | 2212 | 25 | 18 | 1.2 | 1.13-1.27 | | 31 | 19 | 1.25 | 1.17-1.34 | |
| Lower stomach | 6906 | 2050 | 26 | 20 | 1.2 | 1.13-1.28 | | 33 | 22 | 1.25 | 1.17-1.34 | |
| Overlapping | 4649 | 2083 | 13 | 12 | 1.23 | 1.16-1.31 | | 14 | 12 | 1.25 | 1.18-1.34 | |
| Stage | | | | | | | **< 0.001** | | | | | 0.186 |
| Ⅰ | 9550 | 1872 | 68 | 90 | 1.14 | 1.06-1.23 | | | | 1.28 | 1.17-1.39 | |
| Ⅱ | 5194 | 1105 | 34 | 33 | 1.26 | 1.16-1.36 | | 42 | 36 | 1.3 | 1.19-1.42 | |
| Ⅲ | 3715 | 1204 | 21 | 19 | 1.28 | 1.18-1.38 | | 23 | 20 | 1.29 | 1.19-1.4 | |
| Ⅳ | 11898 | 3798 | 9 | 8 | 1.19 | 1.15-1.24 | | 9 | 8 | 1.21 | 1.16-1.26 | |
| Peritoneal cytology[b] | | | | | | | 0.244 | | | | | 0.076 |
| Negative | 16094 | 4081 | 29 | 23 | 1.24 | 1.19-1.29 | | 38 | 26 | 1.3 | 1.25-1.36 | |
| Positive | 5640 | 1857 | 8 | 7 | 1.16 | 1.1-1.23 | | 8 | 7 | 1.17 | 1.11-1.24 | |
| Surgery | | | | | | | 0.057 | | | | | 0.245 |
| No | 12816 | 3382 | 9 | 8 | 1.21 | 1.16-1.27 | | 9 | 8 | 1.23 | 1.18-1.29 | |
| Yes | 17541 | 4597 | 45 | 30 | 1.22 | 1.17-1.27 | | 72 | 34 | 1.27 | 1.21-1.33 | |
| Radiation therapy | | | | | | | 0.894 | | | | | 0.794 |
| No | 20710 | 5797 | 18 | 13 | 1.22 | 1.18-1.27 | | 21 | 14 | 1.27 | 1.22-1.32 | |
| Yes | 9647 | 2182 | 22 | 20 | 1.25 | 1.18-1.32 | | 25 | 21 | 1.28 | 1.21-1.36 | |
| Chemotherapy | | | | | | | **< 0.001** | | | | | **< 0.001** |
| No | 11650 | 2418 | 22 | 15 | 1.12 | 1.06-1.19 | | 33 | 18 | 1.19 | 1.12-1.26 | |
| Yes | 18707 | 5561 | 19 | 15 | 1.28 | 1.23-1.32 | | 20 | 15 | 1.3 | 1.25-1.35 | |

Abbreviation: SEER: Surveillance, Epidemiology, End Results; ref.: referent; HR: hazards ratio; CI: Confidence interval; mOS: median overall survival; mCSS: median cause-specific survival; SRC: signet ring cell carcinoma; API: Asian/Pacific Islander; AIA: American Indian/Alaska Native

[a] Adjusted for age (continuous variables), sex, race, primary site, stage, surgery, radiation therapy and chemotherapy.

[b] Data from 2004 to 2015 for peritoneal metastases were used because data after 2015 did not include information of peritoneal cytology.

Table S4.5. Heterogeneity in the association of survival with clinical characteristics between patients with gastric signet ring cell carcinoma and non-signet ring cell carcinoma after propensity score matching[a] without adjusting age, SEER 2004-2020

| Clinical characristics | N, non-SRC (ref.) | N, SRC | OS | | | | | CSS | | | | |
|---|---|---|---|---|---|---|---|---|---|---|---|---|
| | | | mOS (non-SRC) | mOS (SRC) | HR | 95% CI | $P_{heterogeneity}$ | mCSS (non-SRC) | mCSS (SRC) | HR | 95% CI | $P_{heterogeneity}$ |
| Stage | | | | | | | **< 0.001** | | | | | 0.164 |
| Ⅰ | 1870 | 1872 | 69 | 90 | 0.85 | 0.78-0.93 | | | | 1.07 | 0.96-1.19 | |
| Ⅱ | 1104 | 1105 | 38 | 33 | 1.12 | 1.01-1.25 | | 44 | 36 | 1.18 | 1.05-1.32 | |
| Ⅲ | 1183 | 1199 | 23 | 19 | 1.24 | 1.13-1.35 | | 25 | 20 | 1.27 | 1.15-1.4 | |
| Ⅳ | 3817 | 3798 | 10 | 8 | 1.17 | 1.12-1.23 | | 10 | 8 | 1.19 | 1.14-1.25 | |

Abbreviation: SEER: Surveillance, Epidemiology, End Results; ref.: referent; HR: hazards ratio; CI: Confidence interval; mOS: median overall survival; mCSS: median cause-specific survival; SRC: signet ring cell carcinoma;

API: Asian/Pacific Islander; AIA: American Indian/Alaska Native

[a] Adjusted for propensity score (sex, race, primary site, stage, surgery, radiation therapy and chemotherapy)

Table S4.6. Heterogeneity in the association of survival with clinical characteristics between patients with gastric signet ring cell carcinoma and non-signet ring cell carcinoma after propensity score matching[a] without adjusting age and stage, SEER 2004-2020

| Clinical characristics | N, non-SRC (ref.) | N, SRC | OS | | | | | CSS | | | | |
|---|---|---|---|---|---|---|---|---|---|---|---|---|
| | | | mOS (non-SRC) | mOS (SRC) | HR | 95% CI | $P_{heterogeneity}$ | mCSS (non-SRC) | mCSS (SRC) | HR | 95% CI | $P_{heterogeneity}$ |
| Stage | | | | | | | **< 0.001** | | | | | 0.2 |
| Ⅰ | 2128 | 1872 | 70 | 90 | 0.85 | 0.78-0.93 | | | | 1.09 | 0.98-1.21 | |
| Ⅱ | 1286 | 1105 | 39 | 33 | 1.13 | 1.02-1.25 | | 48 | 36 | 1.21 | 1.08-1.35 | |
| Ⅲ | 1051 | 1204 | 23 | 19 | 1.25 | 1.14-1.38 | | 25 | 20 | 1.29 | 1.16-1.42 | |
| Ⅳ | 3514 | 3798 | 10 | 8 | 1.15 | 1.09-1.2 | | 10 | 8 | 1.16 | 1.11-1.22 | |

Abbreviation: SEER: Surveillance, Epidemiology, End Results; ref.: referent; HR: hazards ratio; CI: Confidence interval; mOS: median overall survival; mCSS: median cause-specific survival; SRC: signet ring cell carcinoma;

API: Asian/Pacific Islander; AIA: American Indian/Alaska Native

[a] Adjusted for propensity score (sex, race, primary site, surgery, radiation therapy and chemotherapy)

Table S4.7. Heterogeneity in the association of survival with clinical characteristics between patients with gastric signet ring cell carcinoma and non-signet ring cell carcinoma after propensity score matching[a] without adjusting age, SEER 2004-2020

| Clinical characristics | N, non-SRC (ref.) | N, SRC | OS | | | | | CSS | | | | |
|---|---|---|---|---|---|---|---|---|---|---|---|---|
| | | | mOS (non-SRC) | mOS (SRC) | Univariable | Multivariable | | mCSS (non-SRC) | mCSS (SRC) | Univariable | Multivariable | |
| | | | | | HR (95% CI) | 95% CI | $P_{heterogeneity}$ | | | HR (95% CI) | HR (95% CI) | $P_{heterogeneity}$ |
| Stage | | | | | | | **< 0.001** | | | | | 0.213 |
| Ⅰ | 1870 | 1872 | 69 | 90 | 0.85(0.78-0.93) | 0.9 (0.82-0.98) | | | | 1.07 (0.96-1.19) | 1.13 (1.01-1.26) | |
| Ⅱ | 1104 | 1105 | 38 | 33 | 1.12 (1.01-1.24) | 1.16 (1.05-1.29) | | 44 | 36 | 1.17 (1.05-1.31) | 1.22 (1.09-1.37) | |
| Ⅲ | 1183 | 1199 | 23 | 19 | 1.23 (1.13-1.35) | 1.25 (1.14-1.37) | | 25 | 20 | 1.27 (1.15-1.39) | 1.28 (1.16-1.41) | |
| Ⅳ | 3817 | 3798 | 10 | 8 | 1.17 (1.12-1.23) | 1.17 (1.12-1.23) | | 10 | 8 | 1.19 (1.13-1.25) | 1.19 (1.13-1.25) | |

Abbreviation: SEER: Surveillance, Epidemiology, End Results; ref.: referent; HR: hazards ratio; CI: Confidence interval; mOS: median overall survival; mCSS: median cause-specific survival; SRC: signet ring cell carcinoma;

API: Asian/Pacific Islander; AIA: American Indian/Alaska Native

[a] Variables included in the propensity score were sex, race, primary site, stage, surgery, radiation therapy and chemotherapy (except peritoneal cytology).

[a] Adjusted for sex, race, primary site, stage, surgery, radiation therapy and chemotherapy.

Table S4.8. Heterogeneity in the association of survival with clinical characteristics between patients with gastric signet ring cell carcinoma and non-signet ring cell carcinoma after propensity score matching[a] without adjusting age and stage, SEER 2004-2020

| Clinical characristics | N, non-SRC (ref.) | N, SRC | OS | | | | | CSS | | | | |
|---|---|---|---|---|---|---|---|---|---|---|---|---|
| | | | mOS (non-SRC) | mOS (SRC) | Univariable | Multivariable | | mCSS (non-SRC) | mCSS (SRC) | Univariable | Multivariable | |
| | | | | | HR (95% CI) | 95% CI | $P_{heterogeneity}$ | | | HR (95% CI) | HR (95% CI) | $P_{heterogeneity}$ |
| Stage | | | | | | | **< 0.001** | | | | | 0.24 |
| Ⅰ | 2128 | 1872 | 70 | 90 | 0.85 (0.78-0.93) | 0.9 (0.83-0.99) | | | | 1.09 (0.98-1.21) | 1.15 (1.03-1.27) | |
| Ⅱ | 1286 | 1105 | 39 | 33 | 1.14 (1.03-1.25) | 1.13 (1.03-1.25) | | 48 | 36 | 1.21 (1.09-1.35) | 1.21 (1.09-1.35) | |
| Ⅲ | 1051 | 1204 | 23 | 19 | 1.25 (1.14-1.38) | 1.28 (1.16-1.4) | | 25 | 20 | 1.28 (1.16-1.42) | 1.31 (1.18-1.45) | |
| Ⅳ | 3514 | 3798 | 10 | 8 | 1.15 (1.09-1.2) | 1.16 (1.1-1.22) | | 10 | 8 | 1.16 (1.11-1.22) | 1.18 (1.12-1.24) | |

Abbreviation: SEER: Surveillance, Epidemiology, End Results; ref.: referent; HR: hazards ratio; CI: Confidence interval; mOS: median overall survival; mCSS: median cause-specific survival; SRC: signet ring cell carcinoma;

API: Asian/Pacific Islander; AIA: American Indian/Alaska Native

[a] Variables included in the propensity score were sex, race, primary site, surgery, radiation therapy and chemotherapy (except peritoneal cytology).

[a] Adjusted for sex, race, primary site, stage, surgery, radiation therapy and chemotherapy.

Table S4.9. Heterogeneity in the association of survival with clinical characteristics between patients with gastric signet ring cell carcinoma and non-signet ring cell carcinoma before propensity score matching without adjusting age, SEER 2004-2020

| Clinical characristics | N, non-SRC (ref.) | N, SRC | OS | | | | | CSS | | | | |
|---|---|---|---|---|---|---|---|---|---|---|---|---|
| | | | mOS (non-SRC) | mOS (SRC) | Univariable | Multivariable | | mCSS (non-SRC) | mCSS (SRC) | Univariable | Multivariable | |
| | | | | | HR (95% CI) | 95% CI | *P-heterogeneity* | | | HR (95% CI) | HR (95% CI) | *P-heterogeneity* |
| Stage | | | | | | | **< 0.001** | | | | | 0.11 |
| Ⅰ | 9550 | 1872 | 68 | 90 | 0.87 (0.81-0.93) | 0.92 (0.85-0.98) | | | | 1.07 (0.98-1.16) | 1.12 (1.03-1.22) | |
| Ⅱ | 5194 | 1105 | 34 | 33 | 1.05 (0.97-1.13) | 1.17 (1.08-1.27) | | 42 | 36 | 1.13 (1.03-1.23) | 1.25 (1.15-1.37) | |
| Ⅲ | 3715 | 1204 | 21 | 19 | 1.13 (1.05-1.21) | 1.2 (1.11-1.29) | | 23 | 20 | 1.17 (1.09-1.26) | 1.24 (1.15-1.35) | |
| Ⅳ | 11898 | 3798 | 9 | 8 | 1.1 (1.06-1.14) | 1.18 (1.13-1.22) | | 9 | 8 | 1.12 (1.08-1.17) | 1.2 (1.15-1.25) | |

Abbreviation: SEER: Surveillance, Epidemiology, End Results; ref.: referent; HR: hazards ratio; CI: Confidence interval; mOS: median overall survival; mCSS: median cause-specific survival; SRC: signet ring cell carcinoma;

API: Asian/Pacific Islander; AIA: American Indian/Alaska Native

[a] Adjusted for sex, race, primary site, stage, surgery, radiation therapy and chemotherapy.

Table S5. Heterogeneity in the association of surgery site with overall survival by stages between SRC and non-SRC gastric cancers in stage Ⅰ and Ⅱ, SEER 2004-2020

| Surgery site | non-SRC, N (%) | SRC, N (%) | Univariable HR (95% CI) | Multivariable[a] HR (95% CI) |
|---|---|---|---|---|
| no surgery | 3187 (21.6) | 665 (22.3) | 1.19 (1.08-1.3) | 1.27 (1.16-1.39) |
| Local tumor destruction or excision, NOS | 733 (5.0) | 38 (1.28) | 1.59 (1-2.53) | 1.75 (1.07-2.87) |
| Gastrectomy, NOS (partial, subtotal, hemi-) | 5600 (38.0) | 1232 (41.4) | 0.8 (0.73-0.88) | 1.09 (0.99-1.2) |
| Near-total or total gastrectomy, NOS | 1262 (8.6) | 394 (13.2) | 0.84 (0.71-1) | 1.09 (0.91-1.3) |
| Gastrectomy, NOS WITH removal of a portion of esophagus | 2954 (20.0) | 426 (14.3) | 1.1 (0.97-1.26) | 1.3 (1.13-1.48) |
| Gastrectomy with a resection in continuity with the resection of other organs, NOS | 903 (6.1) | 203 (6.8) | 0.78 (0.62-0.98) | 0.97 (0.76-1.23) |

Abbreviation: SEER: Surveillance, Epidemiology; SRC: Signet ring cell carcinoma; HR: hazards ratio; CI: Confidence interval

[a]Adjusted for age (as continuous variable), sex, race and primary site.

Table S6.1. Association between subgroup of N, M stage with mortality for patients with gastric adenocarcinoma, SEER 2004-2020.

| Characteristic | T1 (N, %) | | | | T2 (N, %) | | | | T3 (N, %) | | | | T4 (N, %) | | | |
|---|---|---|---|---|---|---|---|---|---|---|---|---|---|---|---|---|
| | non-SRC (ref.) | SRC | Univariable HR (95% CI) | Multivariable[a] HR (95% CI) | non-SRC (ref.) | SRC | Univariable HR (95% CI) | Multivariable[a] HR (95% CI) | non-SRC (ref.) | SRC | Univariable HR (95% CI) | Multivariable[a] HR (95% CI) | non-SRC (ref.) | SRC | Univariable HR (95% CI) | Multivariable[a] HR (95% CI) |
| N0 | 5791 | 1185 | 0.85 (0.78-0.93) | 1.10 (1.01-1.21) | 4103 | 972 | 1.23 (1.13-1.35) | 1.60 (1.45-1.75) | 819 | 306 | 1.25 (1.07-1.46) | 1.36 (1.16-1.61) | 842 | 338 | 1.11 (0.97-1.27) | 1.17 (1.02-1.36) |
| N1 | 1444 | 260 | 0.95 (0.81-1.11) | 1.20 (1.01-1.41) | 5375 | 1083 | 1.12 (1.03-1.20) | 1.27 (1.18-1.38) | 1967 | 544 | 1.16 (1.05-1.29) | 1.25 (1.12-1.40) | 1252 | 393 | 1.11 (0.98-1.25) | 1.17 (1.03-1.32) |
| N2 | 90 | 28 | 0.52 (0.29-0.92) | 0.66 (0.35-1.23) | 1257 | 385 | 0.99 (0.87-1.13) | 1.12 (0.98-1.28) | 881 | 393 | 1.10 (0.97-1.26) | 1.10 (0.96-1.26) | 317 | 134 | 1.11 (0.90-1.38) | 1.12 (0.90-1.40) |
| N3 | 28 | 7 | 0.87 (0.38-2.03) | 1.33 (0.47-3.74) | 386 | 168 | 1.21 (0.99-1.47) | 1.29 (1.05-1.59) | 411 | 285 | 0.93 (0.79-1.10) | 0.95 (0.80-1.12) | 142 | 103 | 1.21 (0.92-1.58) | 1.31 (0.98-1.74) |
| M0 | 5972 | 1183 | 0.77 (0.70-0.85) | 1.15 (1.04-1.26) | 9431 | 2034 | 1.09 (1.03-1.16) | 1.35 (1.27-1.43) | 3225 | 1158 | 1.20 (1.11-1.29) | 1.29 (1.19-1.39) | 1280 | 454 | 1.15 (1.02-1.28) | 1.26 (1.12-1.42) |
| M1 | 1655 | 344 | 1.16 (1.03-1.31) | 1.22 (1.07-1.39) | 2043 | 647 | 1.25 (1.14-1.37) | 1.27 (1.15-1.40) | 935 | 399 | 1.04 (0.92-1.17) | 1.09 (0.96-1.25) | 1562 | 600 | 1.10 (1.00-1.21) | 1.12 (1.01-1.24) |
| -peritoneal cytology negative[b] | 243 | 46 | 0.98 (0.71-1.37) | 1.06 (0.74-1.51) | 328 | 85 | 1.24 (0.97-1.59) | 1.25 (0.97-1.63) | 145 | 38 | 1.00 (0.69-1.45) | 1.15 (0.76-1.73) | 236 | 82 | 0.98 (0.75-1.27) | 1.03 (0.79-1.36) |
| -peritoneal cytology positive[b] | 964 | 214 | 1.23 (1.05-1.42) | 1.29 (1.11-1.52) | 1024 | 357 | 1.18 (1.04-1.33) | 1.21 (1.06-1.38) | 572 | 267 | 0.96 (0.82-1.11) | 1.01 (0.86-1.18) | 1023 | 412 | 1.10 (0.98-1.24) | 1.12 (0.99-1.27) |

Abbreviation: SEER: Surveillance, Epidemiology, End Results; ref.: referent; HR: hazards ratio; CI: Confidence interval; SRC: signet ring cell carcinoma

[a] Adjusted for age (continuous variable), sex, race, primary site.

[b] Data for peritoneal metastases were used from 2004 to 2015 because data after 2015 could not separate out peritoneal cytological positive groups.

Table S6.2. Analysis of the number of regional lymph node metastasis in different stages for patients with gastric signet ring cell carcinoma and non-signet ring cell carcinoma, SEER 2004-2020

| Number of regional lymph node metastases | quartile | Median | quartile | $Z^a$ | $P^a$ |
|---|---|---|---|---|---|
| Stage Ⅰ | | | | | |
| Non-SRC | 0 | 0 | 0 | 0.936 | 0.349 |
| SRC | 0 | 0 | 0 | | |
| Ⅱ | | | | | |
| Non-SRC | 1 | 2 | 6 | 1.539 | 0.124 |
| SRC | 1 | 2 | 4 | | |
| Ⅲ | | | | | |
| Non-SRC | 2 | 6 | 10 | 5.822 | < 0.001 |
| SRC | 3 | 7 | 11 | | |
| Ⅳ | | | | | |
| Non-SRC | 2 | 7 | 17 | 8.4 | < 0.001 |
| SRC | 4 | 12 | 20 | | |

Abbreviation: SEER: Surveillance, Epidemiology, End Results; SRC: signet ring cell carcinoma.

[a] Derived from rank sum test.

Table S6.3. Association between subgroup of N, M stage with mortality for patients with gastric adenocarcinoma, SEER 2004-2020.

| Stage | T1 (N, %) | | | | | | T2 (N, %) | | | | | | T3 (N, %) | | | | | | T4 (N, %) | | | | | |
|---|---|---|---|---|---|---|---|---|---|---|---|---|---|---|---|---|---|---|---|---|---|---|---|---|
| | non-SRC (ref.) | SRC | m-non-SRC (p25, p75) | m-SRC (p25, p75) | $Z^a$ | $P^a$ | non-SRC (ref.) | SRC | m-non-SRC (p25, p75) | m-SRC (p25, p75) | $Z^a$ | $P^a$ | non-SRC (ref.) | SRC | m-non-SRC (p25, p75) | m-SRC (p25, p75) | $Z^a$ | $P^a$ | non-SRC (ref.) | SRC | m-non-SRC (p25, p75) | m-SRC (p25, p75) | $Z^a$ | $P^a$ |
| N | 7627 (100) | 1527 (100) | 0 (0, 0) | 0 (0, 0) | -0.24 | 0.811 | 11480 (100) | 2684 (100) | 1 (0, 4) | 2 (0, 7) | -10.62 | < 0.001 | 4164 (100) | 1560 (100) | 4 (1, 10) | 7 (2, 14.25) | -9.57 | < 0.001 | 2883 (100) | 1074 (100) | 3 (1, 9) | 5 (1, 14) | -4.56 | < 0.001 |
| N0 | 5791 (75.9) | 1185(77.6) | 0 (0, 0) | 0 (0, 0) | -0.83 | 0.407 | 4103 (35.7) | 927 (34.5) | 0 (0, 0) | 0 (0, 0) | -0.38 | 0.703 | 819 (19.7) | 306 (19.6) | 0 (0, 0) | 0 (0, 0) | -0.57 | 0.566 | 842 (29.2) | 338 (31.5) | 0 (0, 0) | 0 (0, 0) | -1.63 | 0.103 |
| N1 | 1444 (18.9) | 260 (17) | 1 (1, 2) | 2 (1, 3) | -3.27 | 0.001 | 5375 (46.8) | 1083 (40.4) | 2 (1, 4) | 2 (1, 4) | -4.21 | < 0.001 | 1967 (47.2) | 544 (34.9) | 2 (1, 4) | 3 (2, 4) | -2.92 | 0.004 | 1252 (43.4) | 393 (36.6) | 2 (1, 4) | 3 (1, 5) | -1.82 | 0.069 |
| N2 | 90 (1.2) | 28 (1.8) | 8 (7, 9) | 9 (8, 11) | -1.38 | 0.167 | 1257 (10.9) | 385 (14.3) | 9 (7, 12) | 10 (8, 12) | -2.69 | 0.007 | 881 (21.2) | 393 (25.2) | 10 (8, 12) | 10 (8, 13) | -2.51 | 0.012 | 317 (11) | 134 (12.5) | 10 (8, 12) | 10.5 (8, 13) | -0.86 | 0.391 |
| N3 | 28 (0.4) | 7 (0.5) | 21 (16, 24) | 21 (20, 21) | 0 | 1 | 386 (3.4) | 168 (6.3) | 21 (17, 25) | 20 (17, 25) | -0.15 | 0.884 | 411 (9.9) | 285 (18.3) | 21 (17, 28) | 21 (18, 27) | -0.32 | 0.75 | 142 (4.9) | 103 (9.6) | 21 (18, 26) | 20 (17, 24.5) | -1.02 | 0.306 |
| M0 | 5972 (78.3) | 1183 (77.5) | | | | < 0.001 | 9431 (82.2) | 2034 (75.8) | | | | < 0.001 | 3225 (77.4) | 1158 (74.2) | | | | <0.001 | 1280 (44.4) | 454 (42.3) | | | | < 0.001 |
| M1 | 1655 (21.7) | 344 (22.5) | | | | < 0.001 | 2043 (17.8) | 647 (24.1) | | | | < 0.001 | 935 (22.5) | 399 (25.6) | | | | <0.001 | 1562 (54.2) | 600 (55.9) | | | | < 0.001 |

Abbreviation: SEER: Surveillance, Epidemiology, End Results; SRC: signet ring cell carcinoma

Abbreviation: SEER: Surveillance, Epidemiology, End Results; SRC: signet ring cell carcinoma; non-SRC: non-signet ring cell carcinoma; m-SRC: median regional lymph node metastasis number of signet ring cell carcinoma; m-non-SRC: median regional lymph node metastasis number of non-signet ring cell carcinoma.

[a] Derived from rank sum test for N stage and $\chi 2$-test for M stage.

Table S7. Baseline characteristics of gastric cancer groups by LRD staging, SEER 2004-2020.

| Clinical Characteristics | All Gastric cancer (N=40364, %) | SRC (N=8362, %) | non-SRC (N=32002, %) | $P^a$ |
|---|---|---|---|---|
| Age, y | | | | |
| Mean (SD) | 64.96(13.39) | 59.89(14.16) | 66.29(12.85) | < 0.001 |
| <65 | 18449(45.7) | 5070(60.6) | 13379(41.8) | |
| >=65 | 21915(54.3) | 3292(39.4) | 18623(58.2) | < 0.001 |
| Sex | | | | < 0.001 |
| Male | 25948(64.3) | 4263(51.0) | 21685(67.8) | |
| Female | 14416(35.7) | 4099(49.0) | 10317(32.2) | |
| Race/ethnicity | | | | |
| White | 19972(49.5) | 3572(42.7) | 16400(51.2) | < 0.001 |
| Black | 4729(11.7) | 892(10.7) | 3837(12.0) | 0.001 |
| API | 6772(16.8) | 1467(17.5) | 5305(16.6) | 0.037 |
| AIA | 362(0.9) | 80(1.0) | 282(0.9) | 0.557 |
| Hispanic | 8529(21.1) | 2351(28.1) | 6178(19.3) | < 0.001 |
| Primary site | | | | |
| Upper stomach | 15274(37.8) | 1789(21.4) | 13485(42.1) | < 0.001 |
| Middle stomach | 8624(21.4) | 2274(27.2) | 6350(19.8) | < 0.001 |
| Lower stomach | 9347(23.2) | 2130(25.5) | 7217(22.6) | < 0.001 |
| Overlapping | 7119(17.6) | 2169(25.9) | 4950(15.5) | < 0.001 |
| Stage | | | | |
| Localized | 10084(25) | 1676(20.0) | 8408(26.3) | < 0.001 |
| Regional | 15915(39.4) | 3378(40.4) | 12537(39.2) | 0.043 |
| Distant | 14365(35.6) | 3308(39.6) | 11057(34.6) | < 0.001 |
| Peritoneal cytology[b] | | | | |
| Negative | 21007(73.7) | 4237(70.0) | 16770(74.8) | |
| Positive | 7497(26.3) | 1857(30.0) | 5640(25.2) | < 0.001 |
| Surgery | | | | |
| No | 17596(43.6) | 3642(43.6) | 13954(43.6) | |
| Yes | 22768(56.4) | 4720(56.4) | 18048(56.4) | 0.945 |
| Radiation therapy | | | | |
| No | 27861(69) | 6062(72.5) | 21799(68.1) | |
| Yes | 12503(31) | 2300(27.5) | 10203(31.9) | < 0.001 |
| Chemotherapy | | | | |
| No | 14868(36.8) | 2547(30.5) | 12321(38.5) | |
| Yes | 25496(63.2) | 5815(69.5) | 19681(61.5) | < 0.001 |

Abbreviation: SEER: Surveillance, Epidemiology, End Results; SRC: signet ring cell carcinoma; SD: standard deviation; API: Asian/Pacific Islander; AIA: American Indian/Alaska Native

[a] Derived from $\chi$2-test for categorical variables, general linear models for continuous variables.

[b] Data for peritoneal metastases were used from 2004 to 2015 because data after 2015 could not separate out peritoneal cytological positive groups.

Table S8. Association between clinical characteristics with mortality for patients with gastric adenocarcinoma by LRD staging, SEER 2004-2020.

| Clinical Characteristics | N | mOS | Overall Survival | | | | | | Cause-specific Survival | | | | | |
|---|---|---|---|---|---|---|---|---|---|---|---|---|---|---|
| | | | Univariable | | | Multivariable[a] | | | Univariable | | | Multivariable[b] | | |
| | | | HR | 95% CI | *P* | HR | 95% CI | *P* | HR | 95% CI | *P* | HR | 95% CI | *P* |
| Age | 40364 | 18 | 1.01 | 1.01-1.01 | < 0.001 | | | | 1.0 | 1.0-1.0 | 0.26 | | | |
| Age, y | | | | | | | | | | | | | | |
| <65 | 18449 | 19 | 1.0 | 1.0 | | 1.0 | 1.0 | | 1.0 | 1.0 | | / | / | |
| >=65 | 21915 | 17 | 1.16 | 1.13-1.18 | < 0.001 | 1.28 | 1.25-1.31 | < 0.001 | 0.99 | 0.97-1.02 | 0.57 | / | / | / |
| Histology | | | | | | | | | | | | | | |
| Gastric non-SRC (ref.) | 32002 | 19 | 1.0 | 1.0 | | 1.0 | 1.0 | | 1.0 | 1.0 | | 1.0 | 1.0 | |
| Gastric SRC | 8362 | 15 | 1.16 | 1.13-1.19 | < 0.001 | 1.24 | 1.2-1.27 | < 0.001 | 1.26 | 1.22-1.29 | < 0.001 | 1.27 | 1.23-1.31 | < 0.001 |
| Sex | | | | | | | | | | | | | | |
| Male (ref.) | 25948 | 18 | 1.0 | 1.0 | | / | / | | 1.0 | 1.0 | | / | / | |
| Female | 14416 | 18 | 1.0 | 0.97-1.02 | 0.774 | / | / | / | 1.0 | 0.97-1.02 | 0.731 | / | / | / |
| Race/ethnicity | | | | | | | | | | | | | | |
| White | 19972 | 17 | 1.0 | 1.0 | | 1.0 | 1.0 | | 1.0 | 1.0 | | 1.0 | 1.0 | |
| Black | 4729 | 17 | 0.99 | 0.95-1.03 | 0.595 | 1.05 | 1.01-1.09 | 0.014 | 0.97 | 0.93-1.01 | 0.162 | 1.0 | 0.96-1.05 | 0.834 |
| API | 6772 | 26 | 0.72 | 0.7-0.74 | < 0.001 | 0.8 | 0.78-0.83 | < 0.001 | 0.73 | 0.71-0.76 | < 0.001 | 0.83 | 0.8-0.86 | < 0.001 |
| AIA | 362 | 13 | 1.15 | 1.02-1.29 | 0.023 | 1.22 | 1.09-1.37 | 0.001 | 1.16 | 1.02-1.32 | 0.02 | 1.19 | 1.04-1.35 | 0.008 |
| Hispanic | 8529 | 17 | 0.94 | 0.92-0.97 | < 0.001 | 0.93 | 0.9-0.96 | < 0.001 | 0.99 | 0.96-1.02 | 0.594 | 0.93 | 0.9-0.96 | < 0.001 |
| Primary site | | | | | | | | | | | | | | |
| Upper stomach | 15274 | 17 | 1.0 | 1.0 | | 1.0 | 1.0 | | 1.0 | 1.0 | | | | |
| Middle stomach | 8624 | 22 | 0.82 | 0.79-0.84 | < 0.001 | 0.98 | 0.94-1.01 | 0.195 | 0.8 | 0.77-0.82 | < 0.001 | 0.98 | 0.94-1.02 | 0.277 |
| Lower stomach | 9347 | 24 | 0.8 | 0.78-0.83 | < 0.001 | 1.03 | 1-1.07 | 0.05 | 0.77 | 0.75-0.8 | < 0.001 | 1.04 | 1-1.08 | 0.033 |
| Overlapping | 7119 | 12 | 1.26 | 1.22-1.31 | < 0.001 | 1.2 | 1.16-1.24 | < 0.001 | 1.29 | 1.25-1.34 | < 0.001 | 1.21 | 1.17-1.25 | < 0.001 |

| | | | | | | | | | | | | | | |
|---|---|---|---|---|---|---|---|---|---|---|---|---|---|---|
| Stage | | | | | | | | | | | | | | |
| Localized | 10084 | 71 | 1.0 | 1.0 | | 1.0 | 1.0 | | 1.0 | 1.0 | | 1.0 | 1.0 | |
| Regional | 15915 | 24 | 1.74 | 1.68-1.79 | < 0.001 | 2.35 | 2.27-2.43 | < 0.001 | 2.29 | 2.2-2.38 | < 0.001 | 3.04 | 2.92-3.18 | < 0.001 |
| Distant | 14365 | 8 | 4.7 | 4.55-4.86 | < 0.001 | 3.74 | 3.6-3.9 | < 0.001 | 6.61 | 6.35-6.87 | < 0.001 | 4.87 | 4.65-5.09 | < 0.001 |
| $P_{\text{trend}}$ | | | 2.22 | 2.18-2.25 | < 0.001 | 1.88 | 1.84-1.91 | < 0.001 | 2.60 | 2.55-2.65 | < 0.001 | 2.07 | 2.02-2.11 | < 0.001 |
| Peritoneal cytology[c] | | | | | | | | | | | | | | |
| Negative | 21007 | 27 | 1.0 | 1.0 | | | | | 1.0 | 1.0 | | | | |
| Positive | 7497 | 7 | 3.29 | 3.2-3.39 | < 0.001 | | | | 3.69 | 3.58-3.81 | < 0.001 | | | |
| Surgery | | | | | | | | | | | | | | |
| No (ref.) | 17596 | 9 | 1.0 | 1.0 | | 1.0 | 1.0 | | 1.0 | 1.0 | | 1.0 | 1.0 | |
| Yes | 22768 | 41 | 0.26 | 0.26-0.27 | < 0.001 | 0.34 | 0.33-0.35 | < 0.001 | 0.23 | 0.23-0.24 | < 0.001 | 0.32 | 0.31-0.33 | < 0.001 |
| Radiation therapy | | | | | | | | | | | | | | |
| No (ref.) | 27861 | 16 | 1.0 | 1.0 | | 1.0 | 1.0 | | 1.0 | 1.0 | | 1.0 | 1.0 | |
| Yes | 12503 | 21 | 0.87 | 0.85-0.89 | < 0.001 | 1.05 | 1.02-1.08 | < 0.001 | 0.88 | 0.86-0.91 | < 0.001 | 1.06 | 1.03-1.09 | < 0.001 |
| Chemotherapy | | | | | | | | | | | | | | |
| No (ref.) | 14868 | 19 | 1.0 | 1.0 | | 1.0 | 1.0 | | 1.0 | 1.0 | | 1.0 | 1.0 | |
| Yes | 25496 | 18 | 1.1 | 1.08-1.13 | < 0.001 | 0.58 | 0.56-0.6 | < 0.001 | 1.26 | 1.23-1.29 | < 0.001 | 0.57 | 0.55-0.59 | < 0.001 |

Abbreviation: SEER: Surveillance, Epidemiology, End Results; ref.: referent; HR: hazards ratio; CI: Confidence interval; mOS: median overall survival; SRC: signet ring cell carcinoma; API: Asian/Pacific Islander; AIA: American Indian/Alaska Native

[a] Variables included in multivariate analysis for overall survival were age (>= 65, < 65 years old), primary site, histology, race, stage, surgery, radiation therapy and chemotherapy.

[b] Variables included in multivariate analysis for cause-specific survival were primary site, histology, race, stage, surgery, radiation therapy and chemotherapy.

[c] Data from 2004 to 2015 for peritoneal metastases were used because data after 2015 did not include information of peritoneal cytology.

Table S9. Baseline characteristics of gastric cancer (signet ring cell carcinoma and non-signet ring cell carcinoma) groups after propensity score matching[a], SEER 2004-2020

| Clinical Characteristics | All Gastric cancer (N=16602, %) | SRC (N=8301, %) | non-SRC (N=8301, %) | $P^a$ |
|---|---|---|---|---|
| OS (months) | | | | < 0.001 |
| CSS (months) | | | | < 0.001 |
| Survival months | 32.18(42.09) | 30.39(41.02) | 33.96(43.06) | <0.001 |
| Age, y | | | | |
| Mean (SD) | 61.02(14.01) | 59.98(14.15) | 62.06(13.79) | < 0.001 |
| <65 | 9985(60.1) | 5009(60.3) | 4976(59.9) | |
| >=65 | 6617(39.9) | 3292(39.7) | 3325(40.1) | 0.612 |
| Sex | | | | 0.744 |
| Male | 8504(51.2) | 4263(51.4) | 4241(51.1) | |
| Female | 8098(48.8) | 4038(48.6) | 4060(48.9) | |
| Race/ethnicity | | | | |
| White | 7093(42.7) | 3559(42.9) | 3534(42.6) | 0.707 |
| Black | 1771(10.7) | 892(10.7) | 879(10.6) | 0.763 |
| API | 2946(17.7) | 1464(17.6) | 1482(17.9) | 0.73 |
| AIA | 146(0.9) | 79(1.0) | 67(0.8) | 0.361 |
| Hispanic | 4646(28) | 2307(27.8) | 2339(28.2) | 0.592 |
| Primary site | | | | |
| Upper stomach | 3585(21.6) | 1789(21.6) | 1796(21.6) | 0.91 |
| Middle stomach | 4508(27.2) | 2247(27.1) | 2261(27.2) | 0.821 |
| Lower stomach | 4256(25.6) | 2130(25.7) | 2126(25.6) | 0.957 |
| Overlapping | 4253(25.6) | 2135(25.7) | 2118(25.5) | 0.776 |
| Stage | | | | |
| Localized | 3329(20.1) | 1673(20.2) | 1656(19.9) | 0.756 |
| Regional | 6770(40.8) | 3368(40.6) | 3402(41.0) | 0.602 |
| Distant | 6503(39.2) | 3260(39.3) | 3243(39.1) | 0.799 |
| Peritoneal cytology[c] | | | | |
| Negative | 8405(69.7) | 4224(69.8) | 4181(69.6) | |
| Positive | 3646(30.3) | 1824(30.2) | 1822(30.4) | 0.833 |
| Surgery | | | | |
| No | 7248(43.7) | 3630(43.7) | 3618(43.6) | |
| Yes | 9354(56.3) | 4671(56.3) | 4683(56.4) | 0.863 |
| Radiation therapy | | | | |
| No | 12026(72.4) | 6012(72.4) | 6014(72.4) | |
| Yes | 4576(27.6) | 2289(27.6) | 2287(27.6) | 0.986 |
| Chemotherapy | | | | |
| No | 5059(30.5) | 2546(30.7) | 2513(30.3) | |
| Yes | 11543(69.5) | 5755(69.3) | 5788(69.7) | 0.59 |

Abbreviation: SEER: Surveillance, Epidemiology, End Results; SRC: signet ring cell carcinoma; API: Asian/Pacific Islander; AIA: American Indian/Alaska Native

[a] Variables included in the propensity score were age (categorical variables), sex, race, primary site, stage, surgery, radiation therapy and chemotherapy.

[b] Derived from $\chi 2$-test for categorical variables, general linear models for continuous variables.

[c] Data for peritoneal metastases were used from 2004 to 2015 because data after 2015 could not separate out peritoneal cytological positive groups.

Table S10. Association between clinical characteristics with survival for patients with gastric adenocarcinoma by LRD staging after propensity score matching[c], SEER 2004-2020.

| Characteristic | N | mOS | Overall Survival | | | | | | Cause-specific Survival | | | | | |
|---|---|---|---|---|---|---|---|---|---|---|---|---|---|---|
| | | | Univariable | | | Multivariable[a] | | | Univariable | | | Multivariable[a] | | |
| | | | HR | 95% CI | *P* | HR | 95% CI | *P* | HR | 95% CI | P | HR | 95% CI | *P* |
| Age | 16602 | 16 | 1.01 | 1.01-1.01 | < 0.001 | | | | 1.0 | 1.0-1.0 | 0.153 | | | |
| Age, y | | | | | | | | | | | | | | |
| < 65 (ref.) | 9985 | 17 | 1.0 | 1.0 | | 1.0 | 1.0 | | 1.0 | 1.0 | | 1.0 | 1.0 | |
| >= 65 | 6617 | 15 | 1.2 | 1.16-1.25 | < 0.001 | 1.29 | 1.24-1.33 | < 0.001 | 1.06 | 1.02-1.1 | 0.002 | 1.15 | 1.11-1.2 | < 0.001 |
| Histology | | | | | | | | | | | | | | |
| Gastric non-SRC (ref.) | 8301 | 19 | 1.0 | 1.0 | | 1.0 | 1.0 | | 1.0 | 1.0 | | 1.0 | 1.0 | |
| Gastric SRC | 8301 | 15 | 1.15 | 1.11-1.19 | < 0.001 | 1.21 | 1.17-1.26 | < 0.001 | 1.2 | 1.15-1.24 | < 0.001 | 1.26 | 1.22-1.31 | < 0.001 |
| Sex | | | | | | | | | | | | | | |
| Male (ref.) | 8504 | 16 | 1.0 | 1.0 | | / | / | | 1.0 | 1.0 | | / | / | |
| Female | 8098 | 17 | 0.98 | 0.95-1.02 | 0.312 | / | / | / | 0.99 | 0.95-1.02 | 0.476 | / | / | / |
| Race/ethnicity | | | | | | | | | | | | | | |
| White | 7093 | 16 | 1.0 | 1.0 | | 1.0 | 1.0 | | 1.0 | 1.0 | | 1.0 | 1.0 | |
| Black | 1771 | 17 | 0.93 | 0.88-0.99 | 0.024 | 1.04 | 0.98-1.11 | 0.169 | 0.93 | 0.87-0.99 | 0.018 | 1.01 | 0.95-1.08 | 0.666 |
| API | 2946 | 22 | 0.74 | 0.7-0.78 | < 0.001 | 0.85 | 0.81-0.9 | < 0.001 | 0.77 | 0.73-0.81 | < 0.001 | 0.88 | 0.83-0.93 | < 0.001 |
| AIA | 146 | 14 | 0.96 | 0.8-1.16 | 0.688 | 1.15 | 0.95-1.39 | 0.148 | 0.99 | 0.81-1.21 | 0.906 | 1.17 | 0.95-1.42 | 0.133 |
| Hispanic | 4646 | 15 | 0.97 | 0.93-1.01 | 0.188 | 0.96 | 0.92-1 | 0.061 | 1.02 | 0.97-1.06 | 0.431 | 0.97 | 0.93-1.02 | 0.269 |
| Primary site | | | | | | | | | | | | | | |
| Upper stomach | 3585 | 15 | 1.0 | 1.0 | | 1.0 | 1.0 | | 1.0 | 1.0 | | 1.0 | 1.0 | |
| Middle stomach | 4508 | 20 | 0.77 | 0.73-0.81 | < 0.001 | 0.97 | 0.92-1.03 | 0.288 | 0.76 | 0.72-0.8 | < 0.001 | 0.97 | 0.92-1.03 | 0.303 |
| Lower stomach | 4256 | 22 | 0.76 | 0.72-0.8 | < 0.001 | 1.02 | 0.97-1.08 | 0.488 | 0.74 | 0.7-0.78 | < 0.001 | 1.02 | 0.96-1.08 | 0.533 |

| | | | | | | | | | | | | | | |
|---|---|---|---|---|---|---|---|---|---|---|---|---|---|---|
| Overlapping | 4253 | 12 | 1.17 | 1.11-1.23 | < 0.001 | 1.18 | 1.12-1.24 | < 0.001 | 1.18 | 1.12-1.24 | < 0.001 | 1.17 | 1.11-1.24 | < 0.001 |
| Stage | | | | | | | | | | | | | | |
| Localized | 3329 | 82 | 1.0 | 1.0 | | 1.0 | 1.0 | | 1.0 | 1.0 | | 1.0 | 1.0 | |
| Regional | 6770 | 24 | 1.88 | 1.77-1.98 | < 0.001 | 2.59 | 2.44-2.76 | < 0.001 | 2.34 | 2.19-2.5 | < 0.001 | 3.22 | 3-3.45 | < 0.001 |
| Distant | 6503 | 8 | 5.29 | 5-5.6 | < 0.001 | 4.29 | 4.02-4.58 | < 0.001 | 6.95 | 6.51-7.43 | < 0.001 | 5.4 | 5.02-5.82 | < 0.001 |
| $P_{trend}$ | | | 2.39 | 2.32-2.45 | < 0.001 | 2.0 | 1.92-2.04 | < 0.001 | 2.70 | 2.62-2.78 | < 0.001 | 2.14 | 2.07-2.22 | < 0.001 |
| Peritoneal metastasis[b] | | | | | | | | | | | | | | |
| Negative | 8405 | 26 | 1.0 | 1.0 | | | | | 1.0 | 1.0 | | | | |
| Positive | 3646 | 7 | 3.28 | 3.15-3.43 | < 0.001 | | | | 3.58 | 3.43-3.75 | < 0.001 | | | |

Table S15. Continued

| | | | | | | | | | | | | | | |
|---|---|---|---|---|---|---|---|---|---|---|---|---|---|---|
| Surgery | | | | | | | | | | | | | | |
| No (ref.) | 7248 | 8 | 1.0 | 1.0 | | 1.0 | 1.0 | | 1.0 | 1.0 | | 1.0 | 1.0 | |
| Yes | 9354 | 35 | 0.27 | 0.26-0.28 | < 0.001 | 0.36 | 0.34-0.38 | < 0.001 | 0.25 | 0.24-0.26 | < 0.001 | 0.35 | 0.33-0.37 | < 0.001 |
| Radiation therapy | | | | | | | | | | | | | | |
| No (ref.) | 12026 | 14 | 1.0 | 1.0 | | 1.0 | 1.0 | | 1.0 | 1.0 | | 1.0 | 1.0 | |
| Yes | 4576 | 22 | 0.77 | 0.74-0.8 | < 0.001 | 1.03 | 0.98-1.08 | 0.212 | 0.76 | 0.73-0.79 | < 0.001 | 1.03 | 0.98-1.08 | 0.263 |
| Chemotherapy | | | | | | | | | | | | | | |
| No (ref.) | 5059 | 16 | 1.0 | 1.0 | | 1.0 | 1.0 | | 1.0 | 1.0 | | 1.0 | 1.0 | |
| Yes | 11543 | 17 | 1.09 | 1.04-1.13 | < 0.001 | 0.61 | 0.58-0.64 | < 0.001 | 1.19 | 1.14-1.24 | < 0.001 | 0.62 | 0.59-0.65 | < 0.001 |

Abbreviation: SEER: Surveillance, Epidemiology, End Results; ref.: referent; HR: hazards ratio; CI: Confidence interval; mOS: median overall survival; SRC: signet ring cell carcinoma; API: Asian/Pacific Islander; AIA: American Indian/Alaska Native

[a] Variables included in multivariate analysis were age (>= 65, < 65 years old), primary site, histology, race, stage, surgery, radiation therapy and chemotherapy.

[b] Data from 2004 to 2015 for peritoneal metastases were used because data after 2015 did not include information of peritoneal cytology.

Table S11.1. Heterogeneity in the association of survival with clinical characteristics between patients with gastric signet ring cell carcinoma and non-signet ring cell carcinoma after propensity score matching[a] by LRD staging, SEER 2004-2020

| Clinical characristics | N, non-SRC (ref.) | N, SRC | OS | | | | | CSS | | | | |
|---|---|---|---|---|---|---|---|---|---|---|---|---|
| | | | mOS (non-SRC) | mOS (SRC) | HR[a] | 95% CI | *P-heterogeneity*[a] | mCSS (non-SRC) | mCSS (SRC) | HR[a] | 95% CI | *P-heterogeneity*[a] |
| Age (as continuous variable) | 8301 | 8301 | 19 | 15 | | | 0.379 | 20 | 16 | | | 0.024 |
| Sex | | | | | | | 0.458 | | | | | 0.395 |
| Male | 4241 | 4263 | 19 | 14 | 1.16 | 1.11-1.22 | | 21 | 15 | 1.21 | 1.15-1.28 | |
| Female | 4060 | 4038 | 18 | 15 | 1.13 | 1.07-1.19 | | 20 | 16 | 1.17 | 1.11-1.23 | |
| Race/ethnicity | | | | | | | 0.932 | | | | | 0.924 |
| White | 3534 | 3559 | 18 | 14 | 1.16 | 1.1-1.23 | | 19 | 15 | 1.2 | 1.13-1.27 | |
| Black | 879 | 892 | 19 | 15 | 1.13 | 1.02-1.26 | | 20 | 17 | 1.18 | 1.05-1.32 | |
| API | 1482 | 1464 | 26 | 20 | 1.11 | 1.01-1.21 | | 29 | 20 | 1.15 | 1.05-1.26 | |
| AIA | 67 | 79 | 13 | 14 | 1.13 | 0.78-1.66 | | 18 | 15 | 1.18 | 0.79-1.77 | |
| Hispanic | 2339 | 2307 | 17 | 13 | 1.16 | 1.08-1.24 | | 18 | 14 | 1.21 | 1.13-1.3 | |
| Primary site | | | | | | | 0.096 | | | | | 0.263 |
| Upper stomach | 1796 | 1789 | 17 | 13 | 1.21 | 1.12-1.3 | | 19 | 14 | 1.24 | 1.15-1.34 | |
| Middle stomach | 2261 | 2247 | 22 | 18 | 1.08 | 1.01-1.16 | | 25 | 19 | 1.13 | 1.05-1.21 | |
| Lower stomach | 2126 | 2130 | 25 | 20 | 1.14 | 1.06-1.22 | | 29 | 21 | 1.19 | 1.1-1.29 | |
| Overlapping | 2118 | 2135 | 13 | 11 | 1.2 | 1.12-1.28 | | 14 | 12 | 1.23 | 1.15-1.32 | |
| Stage | | | | | | | **0.001** | | | | | 0.14 |
| Localized | 1656 | 1673 | 79 | 89 | 1.01 | 0.92-1.11 | | | | 1.18 | 1.05-1.33 | |
| Regional | 3402 | 3368 | 28 | 20 | 1.26 | 1.19-1.33 | | 32 | 22 | 1.31 | 1.24-1.4 | |
| Distant | 3243 | 3260 | 9 | 8 | 1.19 | 1.13-1.25 | | 9 | 8 | 1.2 | 1.14-1.26 | |
| Peritoneal cytology[b] | | | | | | | 0.451 | | | | | 0.324 |
| Negative | 4181 | 4224 | 29 | 22 | 1.14 | 1.09-1.2 | | 36 | 25 | 1.23 | 1.16-1.29 | |
| Positive | 1822 | 1824 | 8 | 7 | 1.17 | 1.09-1.25 | | 8 | 7 | 1.17 | 1.1-1.25 | |
| Surgery | | | | | | | 0.516 | | | | | 0.365 |
| No | 3618 | 3630 | 9 | 8 | 1.18 | 1.13-1.24 | | 9 | 8 | 1.2 | 1.14-1.26 | |
| Yes | 4683 | 4671 | 41 | 31 | 1.17 | 1.11-1.23 | | 55 | 35 | 1.25 | 1.18-1.33 | |
| Radiation therapy | | | | | | | 0.109 | | | | | 0.15 |
| No | 6014 | 6012 | 16 | 13 | 1.12 | 1.08-1.17 | | 17 | 14 | 1.17 | 1.12-1.22 | |
| Yes | 2287 | 2289 | 25 | 20 | 1.22 | 1.14-1.31 | | 29 | 21 | 1.27 | 1.18-1.36 | |
| Chemotherapy | | | | | | | **0.001** | | | | | 0.076 |
| No | 2513 | 2546 | 18 | 14 | 1.04 | 0.97-1.11 | | 24 | 16 | 1.12 | 1.04-1.2 | |
| Yes | 5788 | 5755 | 19 | 15 | 1.21 | 1.16-1.26 | | 20 | 15 | 1.23 | 1.18-1.29 | |

Abbreviation: SEER: Surveillance, Epidemiology, End Results; ref.: referent; HR: hazards ratio; CI: Confidence interval; mOS: median overall survival; mCSS: median cause-specific survival; SRC: signet ring cell carcinoma; API: Asian/Pacific Islander; AIA: American Indian/Alaska Native

[a] Adjusted for propensity score (age, sex, race, primary site, stage, surgery, radiation therapy and chemotherapy), and age was as categorical variables.

[b] Data from 2004 to 2015 for peritoneal metastases were used because data after 2015 did not include information of peritoneal cytology.

Table S11.2. Heterogeneity in the association of survival with clinical characteristics between patients with gastric signet ring cell carcinoma and non-signet ring cell carcinoma after propensity score matching[a] by LRD staging, SEER 2004-2020

| Clinical characristics | N, non-SRC (ref.) | N, SRC | OS | | | | | CSS | | | | |
|---|---|---|---|---|---|---|---|---|---|---|---|---|
| | | | mOS (non-SRC) | mOS (SRC) | HR[b] | 95% CI | $P_{heterogeneity}$[b] | mCSS (non-SRC) | mCSS (SRC) | HR[b] | 95% CI | $P_{heterogeneity}$[b] |
| Age (as continuous variable) | 8362 | 8362 | 17 | 15 | | | 0.11 | 19 | 16 | | | **0.001** |
| Sex | | | | | | | 0.732 | | | | | 0.675 |
| Male | 5399 | 4262 | 18 | 14 | 1.15 | 1.1-1.21 | | 20 | 15 | 1.2 | 1.15-1.26 | |
| Female | 2959 | 4096 | 18 | 15 | 1.14 | 1.07-1.2 | | 20 | 16 | 1.18 | 1.11-1.25 | |
| Race/ethnicity | | | | | | | 0.063 | | | | | 0.109 |
| White | 3269 | 3571 | 17 | 14 | 1.14 | 1.08-1.2 | | 19 | 15 | 1.17 | 1.11-1.24 | |
| Black | 899 | 889 | 19 | 15 | 1.14 | 1.02-1.27 | | 21 | 17 | 1.18 | 1.05-1.32 | |
| API | 2190 | 1467 | 26 | 20 | 1.1 | 1.02-1.19 | | 29 | 20 | 1.15 | 1.06-1.26 | |
| AIA | 119 | 80 | 12 | 14 | 0.95 | 0.68-1.31 | | 13 | 15 | 1.01 | 0.72-1.42 | |
| Hispanic | 1880 | 2350 | 18 | 13 | 1.26 | 1.17-1.35 | | 19 | 14 | 1.3 | 1.21-1.41 | |
| Primary site | | | | | | | **0.002** | | | | | **0.015** |
| Upper stomach | 3185 | 1789 | 19 | 13 | 1.3 | 1.22-1.39 | | 20 | 14 | 1.34 | 1.26-1.44 | |
| Middle stomach | 1772 | 2274 | 23 | 17 | 1.09 | 1.01-1.18 | | 25 | 19 | 1.14 | 1.05-1.23 | |
| Lower stomach | 1943 | 2130 | 24 | 20 | 1.11 | 1.03-1.2 | | 28 | 21 | 1.2 | 1.11-1.3 | |
| Overlapping | 1462 | 2169 | 13 | 12 | 1.16 | 1.08-1.25 | | 13 | 12 | 1.19 | 1.1-1.28 | |
| Stage | | | | | | | **0.08** | | | | | 0.158 |
| Localized | 1912 | 1673 | 93 | 89 | 1.11 | 1.01-1.22 | | | | 1.35 | 1.2-1.51 | |
| Regional | 3288 | 3374 | 27 | 20 | 1.24 | 1.17-1.31 | | 30 | 22 | 1.29 | 1.21-1.37 | |
| Distant | 3154 | 3307 | 9 | 8 | 1.18 | 1.12-1.24 | | 9 | 8 | 1.18 | 1.12-1.25 | |
| Surgery | | | | | | | 0.804 | | | | | 0.059 |
| No | 3584 | 3642 | 9 | 8 | 1.16 | 1.1-1.21 | | 9 | 8 | 1.17 | 1.11-1.23 | |
| Yes | 4769 | 4711 | 42 | 30 | 1.18 | 1.12-1.24 | | 57 | 34 | 1.27 | 1.2-1.34 | |
| Radiation therapy | | | | | | | 0.498 | | | | | 0.698 |
| No | 5965 | 6056 | 16 | 13 | 1.13 | 1.09-1.18 | | 18 | 14 | 1.18 | 1.13-1.23 | |
| Yes | 2388 | 2297 | 24 | 20 | 1.18 | 1.11-1.27 | | 27 | 21 | 1.22 | 1.14-1.31 | |
| Chemotherapy | | | | | | | **< 0.001** | | | | | 0.066 |
| No | 2820 | 2543 | 18 | 14 | 1.04 | 0.98-1.11 | | 23 | 16 | 1.11 | 1.04-1.19 | |
| Yes | 5533 | 5810 | 19 | 15 | 1.21 | 1.16-1.27 | | 20 | 15 | 1.23 | 1.18-1.29 | |

Abbreviation: SEER: Surveillance, Epidemiology, End Results; ref.: referent; HR: hazards ratio; CI: Confidence interval; mOS: median overall survival; mCSS: median cause-specific survival; SRC: signet ring cell carcinoma; API: Asian/Pacific Islander; AIA: American Indian/Alaska Native

[a] When we calculated each variable, we adjusted for propensity score (age, sex, race, primary site, stage, surgery, radiation therapy and chemotherapy, stripping out the calculating variable), and age was treated as categorical variables.

[a] Adjusted for propensity score (age, sex, race, primary site, stage, surgery, radiation therapy and chemotherapy), and age was as categorical variables.

Table S12. Heterogeneity in the association of treatment modality with overall survival by stages between SRC and non-SRC gastric cancers, SEER 2004-2020.

| | non-SRC | SRC | | non-SRC | SRC | | |
|---|---|---|---|---|---|---|---|
| Stage | n | n | HR (95% CI) | n | n | HR (95% CI) | *P-heterogeneity* |
| | Surgery (no) | | | Surgery (yes) | | | |
| Localized | 2856 | 650 | 1.21(1.08-1.35) | 6856 | 1348 | 1.06(0.95-1.18) | < 0.001 |
| Regional | 3221 | 747 | 1.24(1.13-1.37) | 10361 | 2980 | 1.32(1.25-1.39) | 0.419 |
| Distant | 11337 | 3471 | 1.18(1.13-1.24) | 2162 | 816 | 1.27(1.15-1.4) | 0.062 |
| -with peritoneal cytology negative | 1568 | 406 | 1.19(1.04-1.35) | 613 | 169 | 1.27(1.04-1.56) | 0.535 |
| -with peritoneal cytology positive | 5762 | 1993 | 1.13(1.06-1.2) | 1048 | 477 | 1.22(1.07-1.38) | 0.29 |
| | Radiation therapy (no) | | | Radiation therapy (yes) | | | |
| Localized | 7817 | 1644 | 1.28(1.17-1.4) | 1895 | 354 | 1.19(1.02-1.39) | 0.554 |
| Regional | 7024 | 2113 | 1.29(1.21-1.37) | 6558 | 1614 | 1.31(1.22-1.4) | 0.431 |
| Distant | 10422 | 3478 | 1.19(1.14-1.25) | 3077 | 609 | 1.13(1.02-1.25) | 0.938 |
| -with peritoneal cytology negative | 1438 | 416 | 1.2(1.05-1.37) | 743 | 159 | 1.18(0.97-1.45) | 0.795 |
| -with peritoneal cytology positive | 5439 | 2195 | 1.13(1.06-1.2) | 1371 | 275 | 1.19(1.02-1.39) | 0.593 |
| | Chemotherapy (no) | | | Chemotherapy (yes) | | | |
| Localized | 6863 | 1269 | 1.18(1.07-1.3) | 2849 | 729 | 1.4(1.23-1.58) | 0.001 |
| Regional | 3815 | 845 | 1.33(1.22-1.45) | 9767 | 2882 | 1.31(1.24-1.39) | 0.885 |
| Distant | 3486 | 980 | 1.04(0.96-1.14) | 10013 | 3307 | 1.24(1.18-1.3) | < 0.001 |
| -with peritoneal cytology negative | 559 | 122 | 0.94(0.75-1.19) | 1622 | 453 | 1.31(1.15-1.48) | 0.003 |
| -with peritoneal cytology positive | 2029 | 659 | 1.05(0.94-1.17) | 4781 | 1811 | 1.18(1.11-1.27) | 0.055 |

Abbreviation: SEER: Surveillance, Epidemiology; SRC: Signet ring cell carcinoma; HR: hazards ratio; CI: Confidence interval

Adjusted for age (as continuous variable), sex, race and primary site.

Data for peritoneal metastases were used from 2004 to 2015 because data after 2015 could not separate out peritoneal cytological positive groups.

Table S11.3. Heterogeneity in the association of survival with clinical characteristics between patients with gastric signet ring cell carcinoma and non-signet ring cell carcinoma after propensity score matching[a] by LRD staging, SEER 2004-2020.

| Clinical characristics | N, non-SRC (ref.) | N, SRC | OS | | | | | CSS | | | | |
|---|---|---|---|---|---|---|---|---|---|---|---|---|
| | | | mOS (non-SRC) | mOS (SRC) | HR[b] | 95% CI | $P_{-heterogeneity}$[b] | mCSS (non-SRC) | mCSS (SRC) | HR[b] | 95% CI | $P_{-heterogeneity}$[b] |
| Age (as continuous variable) | 8301 | 8301 | 19 | 15 | | | 0.714 | 20 | 16 | | | 0.112 |
| Sex | | | | | | | 0.098 | | | | | 0.06 |
| Male | 4241 | 4263 | 19 | 14 | 1.27 | 1.21-1.33 | | 21 | 15 | 1.31 | 1.25-1.38 | |
| Female | 4060 | 4038 | 18 | 15 | 1.2 | 1.14-1.26 | | 20 | 16 | 1.23 | 1.16-1.29 | |
| Race/ethnicity | | | | | | | 0.812 | | | | | 0.796 |
| White | 3534 | 3559 | 18 | 14 | 1.25 | 1.19-1.32 | | 19 | 15 | 1.29 | 1.22-1.36 | |
| Black | 879 | 892 | 19 | 15 | 1.24 | 1.12-1.39 | | 20 | 17 | 1.28 | 1.14-1.44 | |
| API | 1482 | 1464 | 26 | 20 | 1.19 | 1.09-1.3 | | 29 | 20 | 1.22 | 1.11-1.34 | |
| AIA | 67 | 79 | 13 | 14 | 1.25 | 0.84-1.85 | | 18 | 15 | 1.31 | 0.87-1.99 | |
| Hispanic | 2339 | 2307 | 17 | 13 | 1.21 | 1.13-1.3 | | 18 | 14 | 1.26 | 1.18-1.35 | |
| Primary site | | | | | | | 0.145 | | | | | 0.282 |
| Upper stomach | 1796 | 1789 | 17 | 13 | 1.29 | 1.2-1.39 | | 19 | 14 | 1.33 | 1.23-1.44 | |
| Middle stomach | 2261 | 2247 | 22 | 18 | 1.17 | 1.09-1.25 | | 25 | 19 | 1.2 | 1.11-1.29 | |
| Lower stomach | 2126 | 2130 | 25 | 20 | 1.23 | 1.14-1.32 | | 29 | 21 | 1.28 | 1.18-1.38 | |
| Overlapping | 2118 | 2135 | 13 | 11 | 1.26 | 1.18-1.35 | | 14 | 12 | 1.29 | 1.2-1.38 | |
| Stage | | | | | | | **0.001** | | | | | 0.026 |
| Localized | 1656 | 1673 | 79 | 89 | 1.15 | 1.04-1.26 | | | | 1.3 | 1.15-1.46 | |
| Regional | 3402 | 3368 | 28 | 20 | 1.33 | 1.25-1.41 | | 32 | 22 | 1.37 | 1.29-1.46 | |
| Distant | 3243 | 3260 | 9 | 8 | 1.19 | 1.13-1.25 | | 9 | 8 | 1.2 | 1.14-1.26 | |
| Peritoneal cytology[c] | | | | | | | 0.745 | | | | | 0.06 |
| Negative | 4181 | 4224 | 29 | 22 | 1.25 | 1.19-1.31 | | 36 | 25 | 1.31 | 1.24-1.39 | |
| Positive | 1822 | 1824 | 8 | 7 | 1.16 | 1.09-1.24 | | 8 | 7 | 1.17 | 1.09-1.25 | |
| Surgery | | | | | | | 0.882 | | | | | 0.123 |
| No | 3618 | 3630 | 9 | 8 | 1.2 | 1.14-1.26 | | 9 | 8 | 1.21 | 1.15-1.27 | |
| Yes | 4683 | 4671 | 41 | 31 | 1.28 | 1.21-1.35 | | 55 | 35 | 1.35 | 1.27-1.43 | |
| Radiation therapy | | | | | | | 0.134 | | | | | 0.185 |
| No | 6014 | 6012 | 16 | 13 | 1.21 | 1.16-1.26 | | 17 | 14 | 1.25 | 1.19-1.3 | |
| Yes | 2287 | 2289 | 25 | 20 | 1.28 | 1.2-1.37 | | 29 | 21 | 1.32 | 1.23-1.42 | |
| Chemotherapy | | | | | | | **0.001** | | | | | 0.11 |
| No | 2513 | 2546 | 18 | 14 | 1.14 | 1.07-1.22 | | 24 | 16 | 1.21 | 1.12-1.3 | |
| Yes | 5788 | 5755 | 19 | 15 | 1.27 | 1.22-1.33 | | 20 | 15 | 1.29 | 1.24-1.35 | |

Abbreviation: SEER: Surveillance, Epidemiology, End Results; ref.: referent; HR: hazards ratio; CI: Confidence interval; mOS: median overall survival; mCSS: median cause-specific survival; SRC: signet ring cell carcinoma; API: Asian/Pacific Islander; AIA: American Indian/Alaska Native

[a] Variables included in propensity score were age (categorical variable), sex, race, primary site, stage, surgery, radiation therapy, chemotherapy.

[b] Adjusted for age (continuous variables), sex, race, primary site, stage, surgery, radiation therapy and chemotherapy.

[c] Data from 2004 to 2015 for peritoneal metastases were used because data after 2015 did not include information of peritoneal cytology.

Table S11.4. Heterogeneity in the association of survival with clinical characteristics between patients with gastric signet ring cell carcinoma and non-signet ring cell carcinoma after propensity score matching[a] by LRD staging, SEER 2004-2020.

| Clinical characristics | N, non-SRC (ref.) | N, SRC | OS | | | | | CSS | | | | |
|---|---|---|---|---|---|---|---|---|---|---|---|---|
| | | | mOS (non-SRC) | mOS (SRC) | HR[b] | 95% CI | $P_{heterogeneity}$[b] | mCSS (non-SRC) | mCSS (SRC) | HR[b] | 95% CI | $P_{heterogeneity}$[b] |
| Age (as continuous variable) | 8362 | 8362 | 17 | 15 | | | 0.263 | 19 | 16 | | | 0.691 |
| Age (as categorical variable) | | | | | | | 0.444 | | | | | 0.75 |
| <65 | 3434 | 5070 | 18 | 15 | 1.24 | 1.17-1.3 | | 19 | 16 | 1.27 | 1.2-1.34 | |
| >=65 | 4928 | 3292 | 16 | 14 | 1.24 | 1.18-1.31 | | 19 | 15 | 1.27 | 1.2-1.34 | |
| Sex | | | | | | | 0.068 | | | | | 0.034 |
| Male | 5399 | 4262 | 18 | 14 | 1.27 | 1.22-1.33 | | 20 | 15 | 1.32 | 1.26-1.39 | |
| Female | 2959 | 4096 | 18 | 15 | 1.2 | 1.13-1.27 | | 20 | 16 | 1.23 | 1.16-1.3 | |
| Race/ethnicity | | | | | | | 0.695 | | | | | 0.83 |
| White | 3269 | 3571 | 17 | 14 | 1.26 | 1.19-1.33 | | 19 | 15 | 1.3 | 1.23-1.38 | |
| Black | 899 | 889 | 19 | 15 | 1.29 | 1.16-1.44 | | 21 | 17 | 1.32 | 1.18-1.49 | |
| API | 2190 | 1467 | 26 | 20 | 1.22 | 1.12-1.32 | | 29 | 20 | 1.25 | 1.15-1.37 | |
| AIA | 119 | 80 | 12 | 14 | 1.17 | 0.84-1.64 | | 13 | 15 | 1.25 | 0.88-1.77 | |
| Hispanic | 1880 | 2350 | 18 | 13 | 1.27 | 1.18-1.36 | | 19 | 14 | 1.3 | 1.21-1.41 | |
| Primary site | | | | | | | 0.055 | | | | | 0.179 |
| Upper stomach | 3185 | 1789 | 19 | 13 | 1.33 | 1.25-1.43 | | 20 | 14 | 1.37 | 1.28-1.47 | |
| Middle stomach | 1772 | 2274 | 23 | 17 | 1.18 | 1.1-1.27 | | 25 | 19 | 1.21 | 1.12-1.32 | |
| Lower stomach | 1943 | 2130 | 24 | 20 | 1.22 | 1.13-1.31 | | 28 | 21 | 1.28 | 1.18-1.39 | |
| Overlapping | 1462 | 2169 | 13 | 12 | 1.28 | 1.19-1.38 | | 13 | 12 | 1.31 | 1.21-1.42 | |
| Stage | | | | | | | **0.03** | | | | | **0.021** |
| Localized | 1912 | 1673 | 93 | 89 | 1.18 | 1.07-1.3 | | | | 1.33 | 1.18-1.5 | |
| Regional | 3288 | 3374 | 27 | 20 | 1.32 | 1.25-1.4 | | 30 | 22 | 1.36 | 1.28-1.45 | |
| Distant | 3154 | 3307 | 9 | 8 | 1.19 | 1.13-1.25 | | 9 | 8 | 1.19 | 1.13-1.26 | |
| Peritoneal cytology[c] | | | | | | | 0.745 | | | | | 0.06 |
| Negative | 4181 | 4224 | 29 | 22 | 1.25 | 1.19-1.31 | | 36 | 25 | 1.31 | 1.24-1.39 | |
| Positive | 1822 | 1824 | 8 | 7 | 1.16 | 1.09-1.24 | | 8 | 7 | 1.17 | 1.09-1.25 | |
| Surgery | | | | | | | 0.998 | | | | | 0.122 |
| No | 3584 | 3642 | 9 | 8 | 1.21 | 1.16-1.27 | | 9 | 8 | 1.22 | 1.16-1.29 | |
| Yes | 4769 | 4711 | 42 | 30 | 1.28 | 1.21-1.34 | | 57 | 34 | 1.34 | 1.27-1.42 | |
| Radiation therapy | | | | | | | 0.265 | | | | | 0.356 |
| No | 5965 | 6056 | 16 | 13 | 1.23 | 1.18-1.28 | | 18 | 14 | 1.27 | 1.21-1.32 | |
| Yes | 2388 | 2297 | 24 | 20 | 1.29 | 1.2-1.37 | | 27 | 21 | 1.32 | 1.23-1.42 | |
| Chemotherapy | | | | | | | **0.002** | | | | | 0.149 |
| No | 2820 | 2543 | 18 | 14 | 1.16 | 1.08-1.23 | | 23 | 16 | 1.22 | 1.14-1.31 | |
| Yes | 5533 | 5810 | 19 | 15 | 1.29 | 1.23-1.34 | | 20 | 15 | 1.3 | 1.25-1.36 | |

Abbreviation: SEER: Surveillance, Epidemiology, End Results; ref.: referent; HR: hazards ratio; CI: Confidence interval; mOS: median overall survival; mCSS: median cause-specific survival; SRC: signet ring cell carcinoma; API: Asian/Pacific Islander; AIA: American Indian/Alaska Native

[a] When we calculated each variable, we adjusted for age, sex, race, primary site, stage, surgery, radiation therapy and chemotherapy, stripping out the calculating variable, and age was treated as categorical variables.

[b] Adjusted for age (continuous variables), sex, race, primary site, stage, surgery, radiation therapy and chemotherapy.

Table S11.5. Heterogeneity in the association of survival with clinical characteristics between patients with gastric signet ring cell carcinoma and non-signet ring cell carcinoma by LRD staging, SEER 2004-2020.

| Clinical characristics | N, non-SRC (ref.) | N, SRC | OS | | | | | CSS | | | | |
|---|---|---|---|---|---|---|---|---|---|---|---|---|
| | | | mOS (non-SRC) | mOS (SRC) | HR [a] | 95% CI | $P$-$_{heterogeneity}$ [a] | mCSS (non-SRC) | mCSS (SRC) | HR [a] | 95% CI | $P$-$_{heterogeneity}$ [a] |
| Age (as continuous variable) | 32002 | 8362 | 19 | 15 | | | **< 0.001** | 22 | 16 | | | 0.239 |
| Sex | | | | | | | 0.228 | | | | | 0.084 |
| Male | 21685 | 4263 | 19 | 14 | 1.29 | 1.24-1.34 | | 22 | 15 | 1.34 | 1.29-1.4 | |
| Female | 10317 | 4099 | 19 | 15 | 1.22 | 1.17-1.28 | | 22 | 16 | 1.26 | 1.2-1.32 | |
| Race/ethnicity | | | | | | | 0.468 | | | | | 0.732 |
| White | 16400 | 3572 | 18 | 14 | 1.24 | 1.19-1.3 | | 20 | 15 | 1.29 | 1.23-1.35 | |
| Black | 3837 | 892 | 17 | 15 | 1.27 | 1.16-1.38 | | 20 | 17 | 1.32 | 1.2-1.44 | |
| API | 5305 | 1467 | 29 | 20 | 1.29 | 1.2-1.39 | | 36 | 20 | 1.33 | 1.23-1.44 | |
| AIA | 282 | 80 | 13 | 14 | 1.1 | 0.82-1.47 | | 14 | 15 | 1.16 | 0.85-1.57 | |
| Hispanic | 6178 | 2351 | 19 | 13 | 1.25 | 1.18-1.32 | | 21 | 14 | 1.3 | 1.23-1.39 | |
| Primary site | | | | | | | 0.362 | | | | | 0.944 |
| Upper stomach | 13485 | 1789 | 18 | 13 | 1.3 | 1.23-1.37 | | 20 | 14 | 1.34 | 1.26-1.42 | |
| Middle stomach | 6350 | 2274 | 25 | 17 | 1.2 | 1.13-1.28 | | 30 | 19 | 1.26 | 1.18-1.34 | |
| Lower stomach | 7217 | 2130 | 25 | 20 | 1.25 | 1.18-1.33 | | 32 | 21 | 1.31 | 1.23-1.39 | |
| Overlapping | 4950 | 2169 | 13 | 12 | 1.26 | 1.19-1.34 | | 13 | 12 | 1.29 | 1.21-1.37 | |
| Stage | | | | | | | **< 0.001** | | | | | **0.004** |
| Localized | 8408 | 1676 | 69 | 89 | 1.13 | 1.04-1.22 | | | | 1.26 | 1.15-1.38 | |
| Regional | 12537 | 3378 | 25 | 20 | 1.33 | 1.27-1.39 | | 29 | 22 | 1.38 | 1.31-1.44 | |
| Distant | 11057 | 3308 | 9 | 8 | 1.2 | 1.15-1.26 | | 9 | 8 | 1.22 | 1.16-1.27 | |
| Peritoneal cytology[b] | | | | | | | 0.237 | | | | | 0.099 |
| Negative | 16770 | 4237 | 28 | 22 | 1.26 | 1.21-1.31 | | 36 | 25 | 1.34 | 1.28-1.4 | |
| Positive | 5640 | 1857 | 8 | 7 | 1.16 | 1.1-1.23 | | 8 | 7 | 1.17 | 1.11-1.24 | |
| Surgery | | | | | | | 0.848 | | | | | **0.001** |
| No | 13954 | 3642 | 9 | 8 | 1.22 | 1.17-1.27 | | 9 | 8 | 1.24 | 1.19-1.29 | |
| Yes | 18048 | 4720 | 45 | 30 | 1.29 | 1.23-1.34 | | 72 | 34 | 1.36 | 1.3-1.42 | |
| Radiation therapy | | | | | | | 0.274 | | | | | 0.347 |
| No | 21799 | 6062 | 18 | 13 | 1.25 | 1.21-1.29 | | 20 | 14 | 1.3 | 1.25-1.34 | |
| Yes | 10203 | 2300 | 22 | 20 | 1.28 | 1.21-1.35 | | 24 | 21 | 1.31 | 1.24-1.39 | |
| Chemotherapy | | | | | | | **< 0.001** | | | | | **0.001** |
| No | 12321 | 2547 | 20 | 14 | 1.16 | 1.1-1.22 | | 30 | 16 | 1.23 | 1.16-1.3 | |
| Yes | 19681 | 5815 | 19 | 15 | 1.3 | 1.25-1.34 | | 20 | 15 | 1.32 | 1.27-1.37 | |

Abbreviation: SEER: Surveillance, Epidemiology, End Results; ref.: referent; HR: hazards ratio; CI: Confidence interval; mOS: median overall survival; mCSS: median cause-specific survival; SRC: signet ring cell carcinoma; API: Asian/Pacific Islander; AIA: American Indian/Alaska Native

[a] Adjusted for age (continuous variables), sex, race, primary site, stage, surgery, radiation therapy and chemotherapy.

[b] Data from 2004 to 2015 for peritoneal metastases were used because data after 2015 did not include information of peritoneal cytology

Table S11.6. Heterogeneity in the association of survival with clinical characteristics between patients with gastric signet ring cell carcinoma and non-signet ring cell carcinoma after propensity score matching[a] without adjusting age by LRD staging, SEER 2004-2020

| Clinical characristics | N, non-SRC (ref.) | N, SRC | OS | | | | | CSS | | | | |
|---|---|---|---|---|---|---|---|---|---|---|---|---|
| | | | mOS (non-SRC) | mOS (SRC) | HR | 95% CI | $P_{heterogeneity}$ | mCSS (non-SRC) | mCSS (SRC) | HR | 95% CI | $P_{heterogeneity}$ |
| Stage | | | | | | | **< 0.001** | | | | | **0.009** |
| Localized | 1661 | 1676 | 68 | 89 | 0.86 | 0.78-0.94 | | | | 1.06 | 0.95-1.19 | |
| Regional | 3387 | 3378 | 27 | 20 | 1.23 | 1.16-1.3 | | 32 | 22 | 1.3 | 1.23-1.39 | |
| Distant | 3290 | 3284 | 9 | 8 | 1.18 | 1.12-1.24 | | 9 | 8 | 1.19 | 1.13-1.25 | |

Abbreviation: SEER: Surveillance, Epidemiology, End Results; ref.: referent; HR: hazards ratio; CI: Confidence interval; mOS: median overall survival; mCSS: median cause-specific survival; SRC: signet ring cell carcinoma; API: Asian/Pacific Islander; AIA: American Indian/Alaska Native

[a] Adjusted for propensity score (sex, race, primary site, stage, surgery, radiation therapy and chemotherapy)

Table S11.7. Heterogeneity in the association of survival with clinical characteristics between patients with gastric signet ring cell carcinoma and non-signet ring cell carcinoma after propensity score matching[a] without adjusting age and stage by LRD staging, SEER 2004-2020

| Clinical characristics | N, non-SRC (ref.) | N, SRC | OS | | | | | CSS | | | | |
|---|---|---|---|---|---|---|---|---|---|---|---|---|
| | | | mOS (non-SRC) | mOS (SRC) | HR | 95% CI | $P_{heterogeneity}$ | mCSS (non-SRC) | mCSS (SRC) | HR | 95% CI | $P_{heterogeneity}$ |
| Stage | | | | | | | **< 0.001** | | | | | 0.055 |
| Localized | 1815 | 1676 | 70 | 89 | 0.87 | 0.79-0.95 | | | | 1.09 | 0.97-1.22 | |
| Regional | 3379 | 3378 | 27 | 20 | 1.21 | 1.14-1.28 | | 31 | 22 | 1.28 | 1.21-1.36 | |
| Distant | 3168 | 3308 | 9 | 8 | 1.2 | 1.14-1.26 | | 9 | 8 | 1.21 | 1.15-1.28 | |

Abbreviation: SEER: Surveillance, Epidemiology, End Results; ref.: referent; HR: hazards ratio; CI: Confidence interval; mOS: median overall survival; mCSS: median cause-specific survival; SRC: signet ring cell carcinoma; API: Asian/Pacific Islander; AIA: American Indian/Alaska Native

[a] Adjusted for propensity score (sex, race, primary site, surgery, radiation therapy and chemotherapy)

Table S11.8. Heterogeneity in the association of survival with clinical characteristics between patients with gastric signet ring cell carcinoma and non-signet ring cell carcinoma after propensity score matching[a] without adjusting age by LRD staging, SEER 2004-2020

| Clinical characristics | N, non-SRC (ref.) | N, SRC | OS | | | | | CSS | | | | |
|---|---|---|---|---|---|---|---|---|---|---|---|---|
| | | | mOS (non-SRC) | mOS (SRC) | Univariable | Multivariable | | mCSS (non-SRC) | mCSS (SRC) | Univariable | Multivariable | |
| | | | | | HR (95% CI) | 95% CI | $P_{heterogeneity}$ | | | HR (95% CI) | HR (95% CI) | $P_{heterogeneity}$ |
| Stage | | | | | | | **< 0.001** | | | | | **0.002** |
| Localized | | | | | 0.85 (0.78-0.94) | 0.89 (0.81-0.98) | | | | 1.06 (0.95-1.19) | 1.11 (0.99-1.24) | |
| Regional | 1661 | 1676 | 68 | 89 | 1.22 (1.16-1.3) | 1.26 (1.19-1.33) | | 32 | 22 | 1.3 (1.23-1.38) | 1.34 (1.26-1.43) | |
| Distant | 3387 | 3378 | 27 | 20 | 1.17 (1.12-1.24) | 1.17 (1.11-1.23) | | 9 | 8 | 1.19 (1.13-1.25) | 1.18 (1.12-1.25) | |

Abbreviation: SEER: Surveillance, Epidemiology, End Results; ref.: referent; HR: hazards ratio; CI: Confidence interval; mOS: median overall survival; mCSS: median cause-specific survival; SRC: signet ring cell carcinoma; API: Asian/Pacific Islander; AIA: American Indian/Alaska Native

[a] Variables included in the propensity score were sex, race, primary site, stage, surgery, radiation therapy and chemotherapy (except peritoneal cytology).

[a] Adjusted for sex, race, primary site, stage, surgery, radiation therapy and chemotherapy.

Table S11.9. Heterogeneity in the association of survival with clinical characteristics between patients with gastric signet ring cell carcinoma and non-signet ring cell carcinoma after propensity score matching[a] without adjusting age and stage by LRD staging, SEER 2004-2020

| Clinical characristics | N, non-SRC (ref.) | N, SRC | OS | | | | | CSS | | | | |
|---|---|---|---|---|---|---|---|---|---|---|---|---|
| | | | mOS (non-SRC) | mOS (SRC) | Univariable | Multivariable | | mCSS (non-SRC) | mCSS (SRC) | Univariable | Multivariable | |
| | | | | | HR (95% CI) | 95% CI | $P_{heterogeneity}$ | | | HR (95% CI) | HR (95% CI) | $P_{heterogeneity}$ |
| Stage | | | | | | | **< 0.001** | | | | | **0.011** |

| | | | | | | | | | | | |
|---|---|---|---|---|---|---|---|---|---|---|---|
| Localized | 1815 | 1676 | 70 | 89 | 0.87 (0.79-0.95) | 0.9 (0.82-0.98) | | | | 1.09 (0.97-1.22) | 1..11 (1-1.25) |
| Regional | 3379 | 3378 | 27 | 20 | 1.21 (1.14-1.28) | 1.25 (1.18-1.33) | | 31 | 22 | 1.28 (1.21-1.36) | 1.33 (1.26-1.42) |
| Distant | 3168 | 3308 | 9 | 8 | 1.19 (1.14-1.26) | 1.18 (1.12-1.24) | | 9 | 8 | 1.21 (1.15-1.27) | 1.13 (1.13-1.25) |

Abbreviation: SEER: Surveillance, Epidemiology, End Results; ref.: referent; HR: hazards ratio; CI: Confidence interval; mOS: median overall survival; mCSS: median cause-specific survival; SRC: signet ring cell carcinoma;

API: Asian/Pacific Islander; AIA: American Indian/Alaska Native

[a] Variables included in the propensity score were sex, race, primary site, surgery, radiation therapy and chemotherapy (except peritoneal cytology).

[a] Adjusted for sex, race, primary site, stage, surgery, radiation therapy and chemotherapy.

Table S11.10. Heterogeneity in the association of survival with clinical characteristics between patients with gastric signet ring cell carcinoma and non-signet ring cell carcinoma before propensity score matching without adjusting age by LRD staging, SEER 2004-2020

| Clinical characristics | N, non-SRC (ref.) | N, SRC | OS | | | | | CSS | | | | |
|---|---|---|---|---|---|---|---|---|---|---|---|---|
| | | | mOS (non-SRC) | mOS (SRC) | Univariable | Multivariable | | mCSS (non-SRC) | mCSS (SRC) | Univariable | Multivariable | |
| | | | | | HR (95% CI) | 95% CI | $P_{heterogeneity}$ | | | HR (95% CI) | HR (95% CI) | $P_{heterogeneity}$ |
| Stage | | | | | | | **< 0.001** | | | | | 0.002 |
| Localized | 8408 | 1676 | 69 | 89 | 0.89 (0.82-0.95) | 0.9 (0.84-0.97) | | | | 1.1 (1.01-1.2) | 1.1 (1.01-1.21) | |
| Regional | 12537 | 3378 | 25 | 20 | 1.14 (1.1-1.2) | 1.25 (1.2-1.31) | | 29 | 22 | 1.23 (1.17-1.29) | 1.33 (1.27-1.4) | |
| Distant | 11057 | 3308 | 9 | 8 | 1.16 (1.11-1.21) | 1.19 (1.14-1.24) | | 9 | 8 | 1.18 (1.13-1.23) | 1.21 (1.16-1.26) | |

Abbreviation: SEER: Surveillance, Epidemiology, End Results; ref.: referent; HR: hazards ratio; CI: Confidence interval; mOS: median overall survival; mCSS: median cause-specific survival; SRC: signet ring cell carcinoma;

API: Asian/Pacific Islander; AIA: American Indian/Alaska Native

[a] Adjusted for sex, race, primary site, stage, surgery, radiation therapy and chemotherapy.
